# The efficacy, safety and immunogenicity Nanocovax: results of a randomized, double-blind, placebo-controlled Phase 3 trial

**DOI:** 10.1101/2022.03.22.22272739

**Authors:** Thuy P. Nguyen, Quyet Do, Lan T. Phan, Dang D. Anh, Hiep Khong, Thuong V. Nguyen, Luong V. Hoang, Duc V. Dinh, Hung N. Pham, Men V. Chu, Toan T. Nguyen, Quang D. Pham, Tri M. Le, Tuyen N.T. Trang, Thanh T. Dinh, Thuong V. Vo, Thao T. Vu, Quynh B.P. Nguyen, Vuong T. Phan, Luong V. Nguyen, Giang T. Nguyen, Phong M. Tran, Thuan D. Nghiem, Tien V. Tran, Tien G. Nguyen, Tuynh Q. Tran, Linh T. Nguyen, Anh T. Do, Dung D. Nguyen, Son A. Ho, Viet T. Nguyen, Dung T. Pham, Hieu B. Tran, Son T. Vu, Su X. Hoang, Trung M. Do, Xuan T. Nguyen, Giang Q. Le, Ton Tran, Thang M. Cao, Huy M. Dao, Thao T.T. Nguyen, Uyen Y Doan, Vy T.T. Le, Linh P. Tran, Ngoc M. Nguyen, Ngoc T. Nguyen, Hang T.T. Pham, Quan H. Nguyen, Hieu T. Nguyen, Hang L.K. Nguyen, Nguyen V. Trang, Anh T.L. Nguyen, Linh T. Nguyen, Anh P. Nguyen, Nhung T.H. Trinh, Ly T.K. Le, Van T. B. Tran, Mai T. N. Chu, My H. Phan, My H. Phan, Hoa T. H. Nguyen, Vinh T. Tran, Mai T.N. Tran, Truc T.T. Nguyen, Phat T. Ha, Hieu T. Huynh, Khanh D. Nguyen, Nghia H.T. Duong, Ung T. Thuan, Chung C. Doan, May, Si M. Do

## Abstract

**Background:** Nanocovax is a recombinant severe acute respiratory syndrome coronavirus 2 subunit vaccine composed of full-length prefusion stabilized recombinant SARS-CoV-2 spike glycoproteins (S-2P) and aluminum hydroxide adjuvant. In a Phase 1 and 2 studies, (NCT04683484) the vaccine was found to be safe and induce a robust immune response in healthy adult participants.

**Methods:** We conducted a multicenter, randomized, double-blind, placebo-controlled study to evaluate the safety, immunogenicity, and protective efficacy of the Nanocovax vaccine against Covid-19 in approximately 13,007 volunteers aged 18 years and over. The immunogenicity was assessed based on Anti-S IgG antibody response, surrogate virus neutralization, wild-type SARS-CoV-2 neutralization and the types of helper T-cell response by intracellular staining (ICS) for interferon gamma (IFNg) and interleukin-4 (IL-4). The vaccine efficacy (VE) was calculated basing on serologically confirmed cases of Covid-19.

**Findings:** Up to day 180, incidences of solicited and unsolicited adverse events (AE) were similar between vaccine and placebo groups. 100 serious adverse events (SAE) were observed in both vaccine and placebo groups (out of total 13007 participants). 96 out of these 100 SAEs were determined to be unrelated to the investigational products. 4 SAEs were possibly related, as determined by the Data and Safety Monitoring Board (DSMB) and investigators. Reactogenicity was absent or mild in the majority of participants and of short duration. These findings highlight the excellent safety profile of Nanocovax.

Regarding immunogenicity, Nanocovax induced robust IgG and neutralizing antibody responses. Importantly, Anti S-IgG levels and neutralizing antibody titers on day 42 were higher than those of natural infected cases. Nanocovax was found to induce Th2 polarization rather than Th1.

Post-hoc analysis showed that the VE against symptomatic disease was 51.5% (95% confidence interval [CI] was [34.4%-64.1%]. VE against severe illness and death were 93.3% [62.2-98.1]. Notably, the dominant strain during the period of this study was Delta variant.

**Interpretation:** Nanocovax 25 microgram (mcg) was found to be safe with the efficacy against symptomatic infection of Delta variant of 51.5%.

**Funding:** Research was funded by Nanogen Pharmaceutical Biotechnology JSC., and the Ministry of Science and Technology of Vietnam; ClinicalTrials.gov number, NCT04922788.

## 1. Introduction

Nanocovax is a subunit vaccine, developed and manufactured at Nanogen Pharmaceutical Biotechnology JSC., containing the extracellular domain of prefusion stabilized recombinant SARS-CoV-2 S glycoproteins bound to aluminum hydroxide adjuvant. In rodent and monkey models, Nanocovax induced high levels of Anti-S antibody (Ab)^1^. Neutralizing antibody titers were evaluated by micro-neutralization assay on the original strain as well as the Delta variant.

In a previous Phase 1 and 2 studies, Nanocovax was found to be safe and induced robust immune responses^2^. Here we report the findings of a randomized, double-blind, placebo-controlled Phase 3 trial started in June 2021, to evaluate the safety, immunogenicity and the VE of 25 mcg recombinant SARS-CoV-2 S glycoprotein in an aluminum adjuvant (0.5 mg/dose) in adults of at least 18 years of age.

## 2. Method

### 2.1. Trial design and oversight

The Phase 3 trial was conducted by the Military Medical University (Ha Noi) and the Pasteur Institute in Ho Chi Minh City, at 4 study sites in Vietnam including Hung Yen, Long An, Tien Giang provinces and Ha Noi. This was a multicenter, randomized, double-blind, placebo-controlled clinical trial to evaluate the safety, immunogenicity, and protective efficacy of the Nanocovax vaccine on Vietnamese volunteers from 18 years of age and older. Eligible participants were men and non-pregnant women, at least 18 years of age with a body mass index (BMI) of 16 to 41 (kg/m^2^). The participants were stratified into 3 age groups: from 18 to 45 years old, from 46 to 60 years old and over 60 years old (Table 1). The group of over 60 years olds accounted for at least 1,200 participants.

**Table 1.**
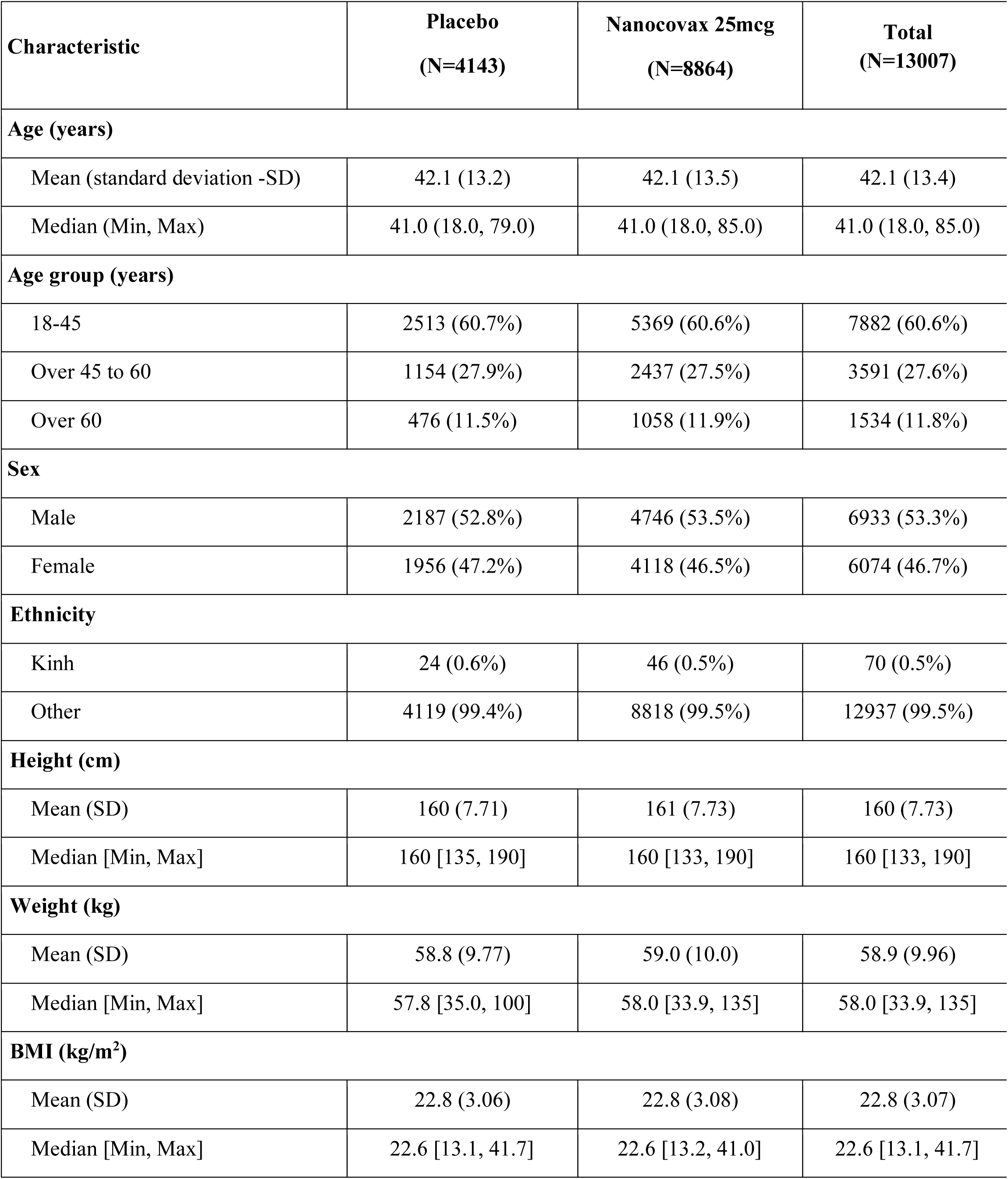
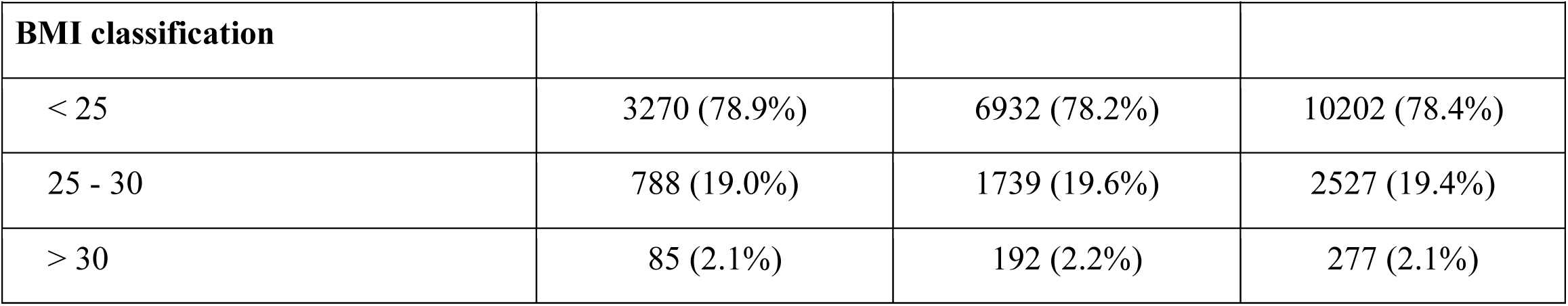
Demographic characteristics of trial population in Phase 3.

13,000 participants were recruited. A subset of the first 1000 participants were evaluated for immunogenicity, in addition to the safety and efficacy (Phase 3a) while the remaining 12,000 participants were followed to evaluate the safety and efficacy of the Nanocovax vaccine (Phase 3b). After enrollment, the first 1000 participants were randomly assigned to either the ‘vaccinate’ or ‘placebo’ groups by the Electronic Clinical Data Management (ECDM) system with a ratio of 6 vaccine:1 placebo. The remaining 12,000 were randomly assigned by the ECDM system to either the ‘vaccine’ or ‘placebo’ group, in a block of 3 with a ratio of 2 vaccine:1 placebo. All participants received 2 injections of vaccine or placebo on Day 0 and Day 28. Trial staff responsible for the vaccine/placebo administration, as well as participants were unaware of vaccine/placebo assignment. Randomization lists, using block randomization stratified by the study group and study site, were generated by the Electronic Data Capture.

All participants were screened by their medical history, clinical and biological examinations, sampling and laboratory tests (complete blood count, biochemistry, urine analysis, testing pregnancy and diagnostic imaging). Participants with a history of Covid-19 or positive results for SARS-CoV-2 at screening period confirmed by real-time reverse transcriptase polymerase-chain-reaction (RT-PCR) were excluded from the trial. All participants signed a written consent before being enrolled in the trial.

Date cut-off for efficacy analysis was December 13^th^, 2021. Intention-to-treat analysis population included participants who had received a first injection. Per-protocol-analysis population included participants receiving 2 injections and those who contracted Covid-19 at least 14 days after the second injection. The safety assessment was conducted on the intention-to-treat population and the immunogenicity assessment was conducted on a subgroup of 1007 participants.

The trials were designed and funded by Nanogen Pharmaceutical Biotechnology JSC and the Ministry of Science and Technology (MOST) of Vietnam. The trial protocol was approved by the Ethics Committee/Protocol Review Board of the Ministry of Health (Vietnam) and was performed in accordance with the ICH-GCP good clinical practice guidelines, with an ethical policy consistent with the “Declaration of Helsinki” and applicable Vietnamese laws and regulations. The authors take responsibility for the data integrity and the fidelity of the trial to the clinical trial protocol.

### 2.2. Trial vaccine, adjuvant, and placebo

The recombinant SARS-CoV-2 spike (S) glycoprotein in Nanocovax were constructed with two proline substitutions (K986P and V987P) for stabilized pre-fusion conformation (S-2). The production of the full-length (including the transmembrane domain) recombinant S protein was optimized in a Chinese Hamster Ovary (CHO) cell-expression system. Clinical grade aluminum hydroxide was manufactured by Croda (Denmark). Recombinant SARS-CoV-2 S protein was absorbed to aluminum adjuvant under mild mixing conditions for 18 hours at 2°C to 8°C. The placebo dose was 0.5 mg of sterile aluminum hydroxide.

### 2.3. Safety assessments

On-site safety follow-up time was 60 minutes after each injection. Follow-up visits to evaluate safety were scheduled on days 28, 42, 178 and 358 after vaccination (Table S1). Participants received instructions for self-monitoring and reporting adverse events within 7 days of each vaccination, as facilitated by the use of a diary with pre-defined reactogenicity criteria. Pre-defined local (injection site) reactogenicity included pain, tenderness, erythema and swelling. Pre-defined systemic reactogenicity included fever, nausea or vomiting, headache, fatigue, malaise, myalgia and arthralgia.

The primary safety outcomes were the number and percentage of participants with solicited local and systemic adverse events which occurred within 7 days of vaccination and laboratory results (serum biochemistry and hematology) at days 0, 7, 28 and 42 according to FDA toxicity scoring^3^. Secondary safety outcomes were the number and percentage of participants with unsolicited events.

AE/SAEs were recorded and evaluated based on the Common Terminology Criteria for Adverse Events 5·0 (CTCAE v5·0) and Guidelines for Toxicity Grading in Healthy Volunteers in the FDA’s Preventive Vaccine Clinical Trials^3,4^. The procedures for recording and evaluation took place continuously from the time of receipt of the first dose to the end of the last visit for each volunteer. Adverse events were assessed in terms of severity score (mild, moderate, severe, potentially life-threatening, or fatal) and relatedness to the vaccine and placebo. Vital sign measurements were assessed according to the FDA toxicity scoring after vaccination^3^. In addition, participants had nasopharyngeal swab tests for SARS-CoV-2 on the day of screening (Day 0, before the first injection), Day 28 (before the second injection) and any time that they developed symptoms associated with SARS-CoV-2 infection.

### 2.4. Immunogenicity assessments

All immunological assays were performed at the National Institute of Hygiene and Epidemiology of Vietnam (NIHE). The primary endpoint was anti-S IgG responses to Nanocovax evaluated by a chemiluminescence immunoassay (CLIA) (Siemens ADVIA Centaur SARS-CoV-2 IgG kit, 11207376). Secondary outcomes were neutralizing antibody titer evaluated by a 50 percent (50%) plaque reduction neutralization test (PRNT_50_) using the original (Wuhan) strain, Alpha variant (B.1.1.7) and Delta variant (B.1.617.2), a competitive enzyme-linked immunosorbent assay (ELISA) based on a surrogate virus neutralization test (sVNT) (Genscript’s cPass™ SARS-CoV-2 Neutralization Antibody Detection Kit, L00847-C) and the T cell response by intracellular cytokine-staining (ICS). The IgG and neutralizing antibody levels of vaccine groups on Day 42 were compared with convalescent sera from symptomatic Covid-19 patients at the Pasteur Institute at Ho Chi Minh City and NIHE, Vietnam. Details of immunological assays are provided in the supplementary appendix.

### 2.5. Vaccine efficacy assessment

The first primary endpoint was vaccine efficacy (VE) against severe illness and death. The second primary endpoint was the VE against symptomatic Covid-19 cases with clinical symptoms with onset at least 14 days after the second dose and virologically confirmed results by a RT-PCR test of nasopharyngeal secretions. Symptomatic Covid-19 was defined according to the criteria of the FDA.

The efficacy was assumed to be at least 50%. To determine whether VE was consistent across different subgroups, VE was estimated in each of the following categories, such as age group (from 18 to 45 years old, from 46 to 60 years old and over 60 years old), gender, and protection against severe illness.

### 2.6. Statistical analysis

The safety was assessed in all participants. Descriptive summary data of participants with solicited/unsolicited systemic and local adverse events (AE), serious adverse events (SAE), medically attended AE and particular events of interests were reported as counts and percentages. For an adverse event that occurred more than once, the analysis was based on only the most severe occurrence and the cause of the event. All serious adverse events were summarized separately. Geometric means (of anti-S IgG concentration and neutralizing antibody titer) and associated 95% confidence intervals (CI) were calculated based on log-transformed data.

The required number of symptomatic Covid-19 events for the primary analysis was at least 69. The primary efficacy end-point was assessed in the per-protocol population and intention to treat population. The VE was defined as the percentage reduction in the hazard ratio between the vaccine and the placebo groups.

The sample size of the Phase 3 study was calculated to test the null hypothesis (H_0_) VE < 50%. The total number of research subjects selected in the study was expected to be 13,000 participants, with the statistical power 80% and 2 interim analysis (IA) at 35% and 70% of the total target cases using the one-sided O’Brien-Fleming boundary method, assuming the VE was 75%, and the alpha error in the one-sided test was 0.025 in the end-of-term analysis. The study also estimated the percentage of participant excluded from the per-protocol population to be 4% and the attack rate in the placebo group to be 0.1%. VE was defined as the percentage reduction in risk for the primary endpoint (Nanocovax 25 mcg vs placebo). A stratified Cox proportional hazards model was used to evaluate the efficacy of the vaccine versus the placebo.

## 3. Result

### 3.1. Trial population

The study was started on June 11, 2021. Of the 13248 participants screened, 13,007 were recruited and randomized: 8861 received Nanocovax 25 mcg and 4146 received placebo. Participants were stratified into 3 age groups: from over 18 to 45 years old, from 46 to 60 years old, and over 60 years old (Figure 1). Demographic characteristics of participants in the Phase 3 study are shown in Table 1. Coexisting conditions of participants were described in table S2.

**Figure 1.**
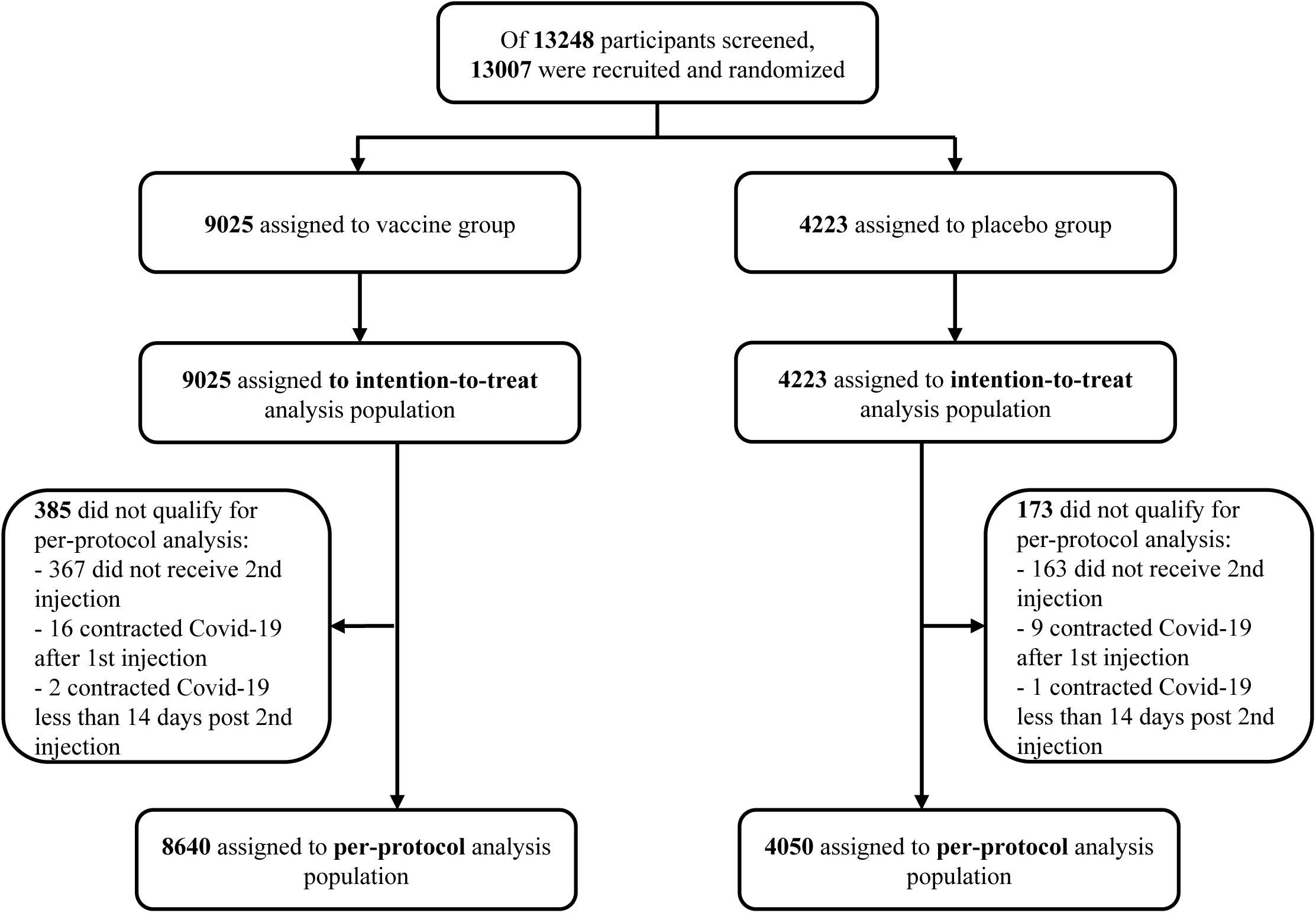
Randomization and analysis populations. Date cutoff for analysis was December 21, 2021. Intention-to-treat analysis population included participants received 1^st^ injection. Per-protocol-analysis population included participants receiving 2 injections and those who contracted Covid-19 at least 14 days after the 2nd injection. Safety assessment was conducted on intention-to-treat population. Immunogenicity assessment was conducted on subgroup of 1007 participants.

### 3.2. Safety outcomes

The incidence of solicited local adverse events in the 7 days after each injection of vaccine and placebo groups were similar (Figure 2). After the first injection, the incidence of pain at the injection site of the vaccine versus the placebo was 40.6 % versus 40.0 %: tenderness: 26.8% versus 26.1%, swelling at the injection site: 0.7% versus 0.8%, redness at the injection site: 0.9% versus 0.9% and pruritus at the injection site: 4.7% versus 5%. Frequencies of solicited local adverse events after dose 2 were lower than those of dose 1 and were similar in both the Nanocovax 25mcg and placebo groups: pain at the injection site 30.3% versus 26.4% (465/1,764), tenderness 18% versus 13.5%, swelling at the injection site was 0.7% versus 0.4%, redness at the injection site 0.6% versus 0.2% and itching 4.7% versus 5.1%. After the second injection, the incidence of local pain after vaccine and placebo was 29.6% versus 27.1%, redness: 0.5% versus 0.2%, itching: 5.1% versus 3.3%, tenderness: 17.9% versus 15.2% and swelling/firmness: 0.5% versus 0.3%. Details of solicited local AE after the first and second injections are shown in Table S3.

**Figure 2.**
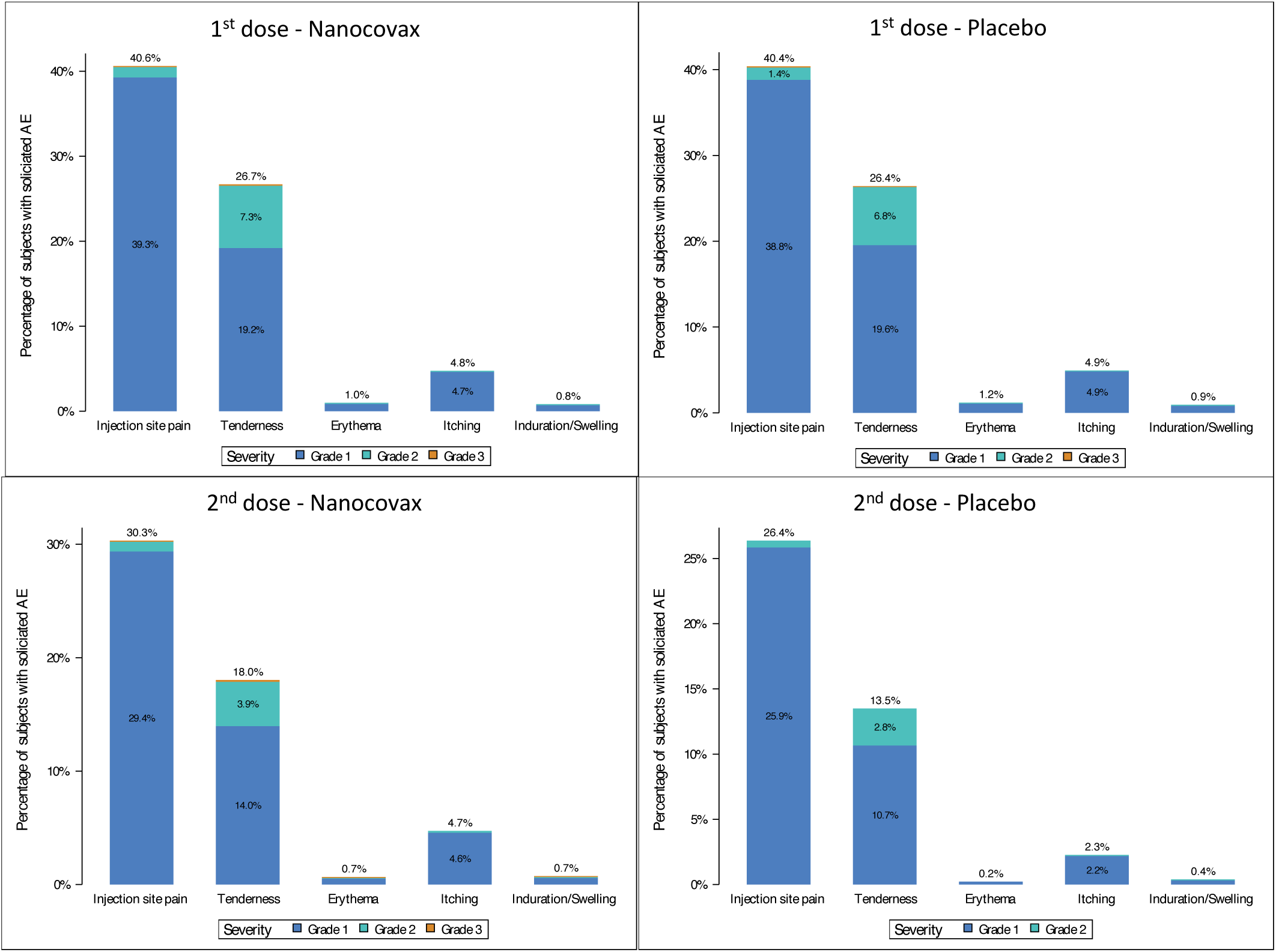
Solicited local adverse events within 7 days after the first (top) and second (bottom) injections.

Incidence of solicited systemic adverse events of the vaccine versus the placebo was similar (Figure 2). 7 days after the first injection the incidence of common systemic AE in vaccine and placebo groups were: fatigue 23.6% versus 24.3%, headache: 16% versus 17.2%, myalgia: 15.6% versus 17.6%, arthralgia: 10% versus 10.7%, nausea/vomiting: 2.9% versus 2.8%, diarrhea: 3.1% versus 3.3% and fever 3% versus 3.4%. All solicited systemic AEs that occurred after the first injection were reversible without sequelae.

Solicited systemic AE 7 days after the second injection were similar between the vaccine and placebo groups (Figure 3). The most common solicited AE were (vaccine versus placebo): fatigue 15.4% versus 13%, headache 7.6% versus 7.4%, myalgia 9.2% versus 8.8%, arthralgia 5.9% versus 4.5%, nausea/vomiting 1.1% versus 1.2%, diarrhea 2% versus 2.3 and fever 1.6% (65/4,021) versus 1.4%. Exact numbers of solicited systemic AEs are shown in Table S4.

**Figure 3.**
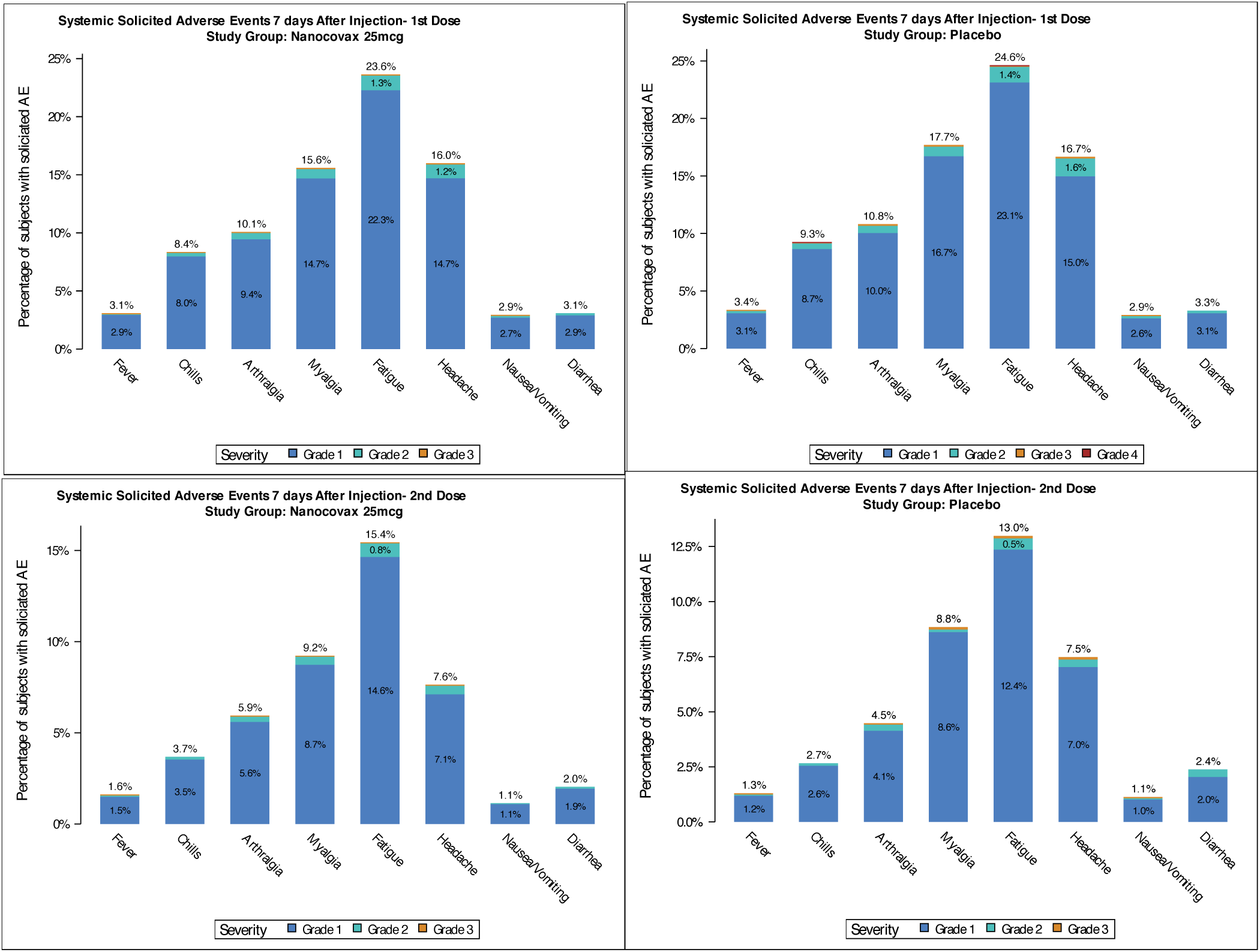
Solicited systemic adverse event occurred within 7 days after the first (top) and second (bottom) injections.

The incidence of unsolicited adverse events was similar among vaccine and placebo groups: 1.68% versus 1.69%, respectively, including hypotension, vestibular disorders, knee pain, hand pain, hyperthyroidism and upper respiratory tract inflammation. Almost all of the unsolicited adverse events were found not to be related to the investigational product and the majority of unsolicited (non-SAE) adverse events in both vaccine and place groups were mild and moderate (Table S5).

There were 100 serious adverse events (SAEs) (including Covid-19 cases) in both study groups (vaccine and placebo) in the trial, of which 96/100 SAEs were assessed as unrelated to the investigational product. Four SAEs were determined to be possibly related by the DSMB and investigator assessments, including Grade 2 anaphylactic reaction, Grade 2 allergic reactions, Grade 3 hypertension and Exacerbation of Chronic Obstructive Pulmonary Disease (COPD). The participant who experienced Grade 2 allergic reactions was in the placebo group. The participants who experienced Grade 2 anaphylactic reaction, Grade 3 hypertension and COPD were in the vaccine group. All participants recovered completely.

### 3.3. Immunogenicity outcomes

Geometric mean concentration (GMC) of anti-S IgG binding antibody units (BAU/mL) and the associated confidence intervals (CI) 95% were shown. Before the first injection, Anti-S IgG levels in the vaccine and placebo groups were 5.7 (CI95% [5.6-5.8]) and 6.1[5.6-6.6], respectively. On Day 42, anti-S IgG concentrations in the vaccine group reached 1254.7[1142.8-1377.6], while that of placebo group remained unchanged 5.9 [5.5-6.4] (Figure 4A). IgG response was negatively correlated with age (Figure S1). GMC Anti-S IgG of 18-45, 46-60 and above 60 year old age groups on Day 42 were 1525.8 [1363.1-1707.8], 983.9 [806.8-1199.9] and 900.7 [708.1-1166.4], respectively (Figure S1A). The correlation coefficient r was -0.2 (Figure S1B).

**Figure 4.**
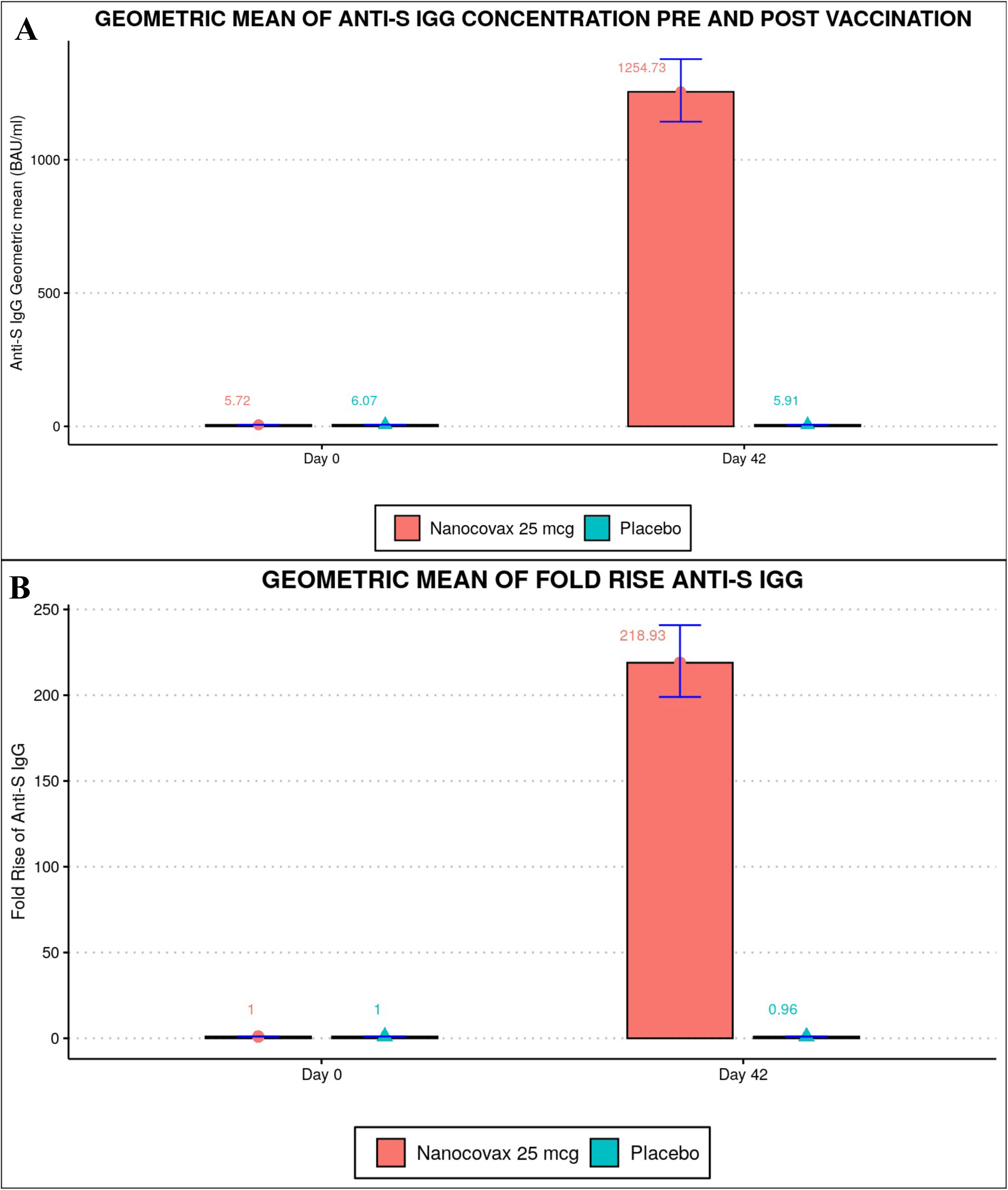
Anti-S IgG responses. A) Geometric mean concentration (GMC) and B) Geometric fold rise (GMFR) of anti-S IgG on day 0 and day 42 of the vaccine (n=858) and placebo (n=145) groups.

Geometric mean fold rise (GMFR) of anti-S IgG was defined as the fold increase in GMC of a given timepoint compared to baseline GMC value of the same group on Day 0. On Day 42, the GMFR of the vaccine group was 218.9 [199-240.9], whilst the GMFR of the placebo group was only 0.96 [0·92-1·11] (Figure 4B). The vaccine group aged from 18-45 years old had the highest GMFR (269.7), followed by 46-60 year old age group (166.4) and those above 60 years (157.9) (Figure S2). The seroconversion rate was defined as GMFR **≥** 4. On Day 42, the seroconversion rate of the vaccine group was 100% (Figure S3).

Neutralizing antibody was qualitatively and quantitatively evaluated using a surrogate virus neutralization test (sVNT) and plaque reduction neutralization test with serum dilution reducing number of plaques by 50% or higher (PRNT_50_), respectively. sVNT results were reported as the percentage (%) of participants in each group positive for the assay. On Day 0, 0.6% and 3.3% participants in the vaccine and placebo groups were positive for sVNT, respectively. On Day 42, 96.4% in vaccine group was positive but only 1.2% positive in placebo group (Figure S4).

Neutralizing antibody, assessed by PRNT_50_, was expressed as the geometric mean titer (GMT) with associated 95% CI. A number of serum samples from vaccine and placebo groups on Day 42 were randomly selected to assess neutralizing antibody titer against the original (Wuhan) strain, the Alpha variant and the Delta variant. GMTs of neutralizing antibody on the Wuhan, Alpha and Delta strains were 57 [46.1-70.5], 36.2 [26.7-49.2] and 29.6 [21.0-41.8], respectively (Figure S5). Meanwhile, the GMT of neutralizing antibody in the placebo group was 10 (half of lower limit of detection).

Type 1 and Type 2 T helper cell (Th1/Th2) response on 77 randomly selected participants (66 in the vaccine group and 11 in the placebo group) were evaluated by ICS using IFNg and IL-4 markers. Although IL-4 and IFNg signals were observed on Day 42, IL-4 signal was more pronounced, suggesting aTh2 polarization induction by Nanocovax (Figure S6). This observation is consistent with the nature of aluminum adjuvants which has been well established for Th2 response induction^5^.

### 3.4. Vaccine Efficacy

The VE was calculated based on per-protocol population of Phase 2 and 3 (n = 12,694) with follow-up times of 180 days (for Phase 3) and 290 days (for Phase 2). The combined VE against symptomatic infection for Phase 2 and 3 was 52.2% [36.0-64.3], which is calculated based on number of cases of symptomatic Covid-19 regardless of the illness severity, with an onset at least 14 days after the second dose occurred in 93 vaccine recipients (24.2 per 1000 person-years) and in 89 placebo recipients (49.9 per 1000 person-years) for a VE adjusted by person-years of 51.5% (95%CI [34.4-64.1]) (Figure 5 and Figure 6).

**Figure 5.**
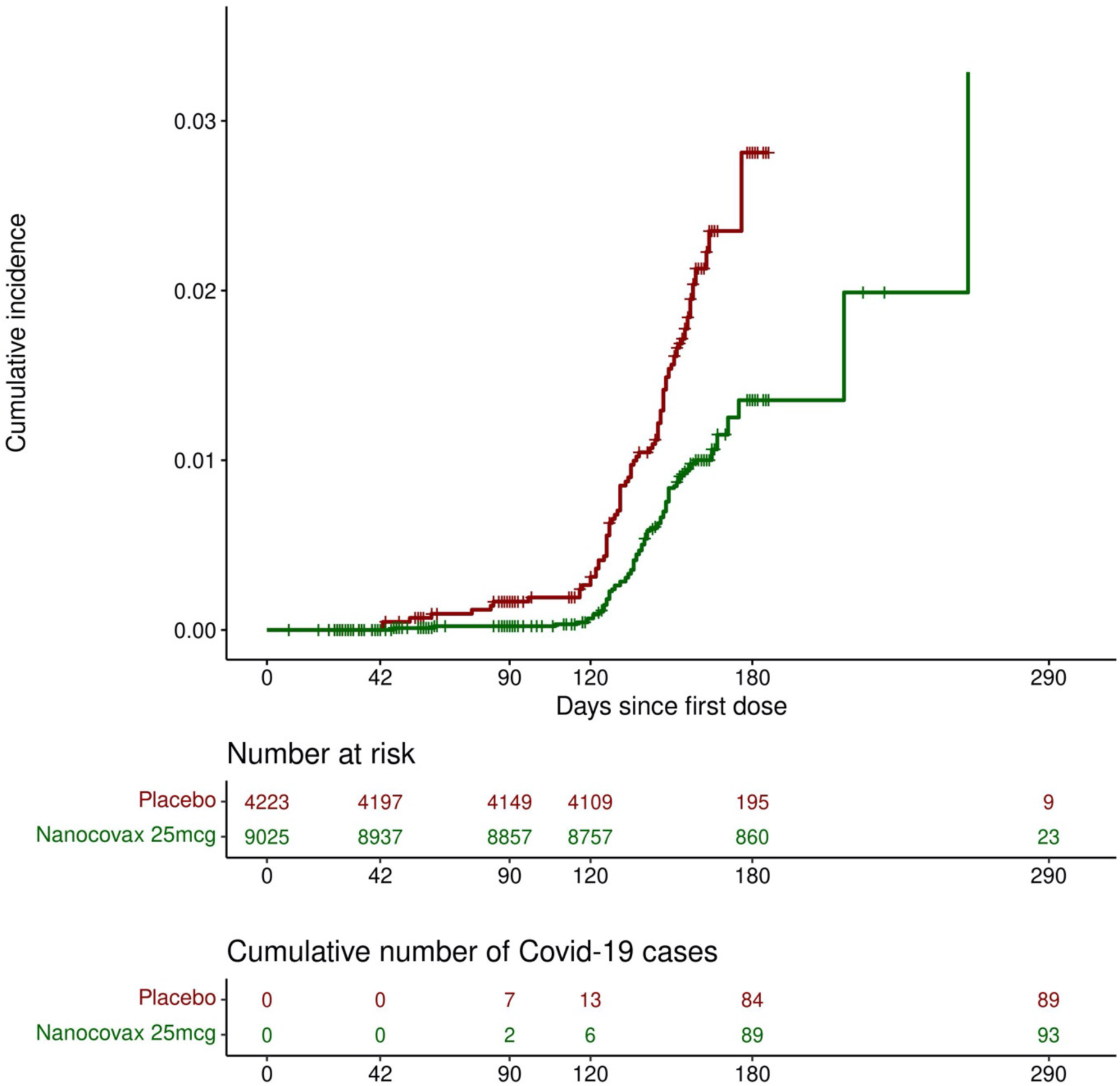
Cumulative incidence of Covid-19 (days since the first dose).

**Figure 6.**
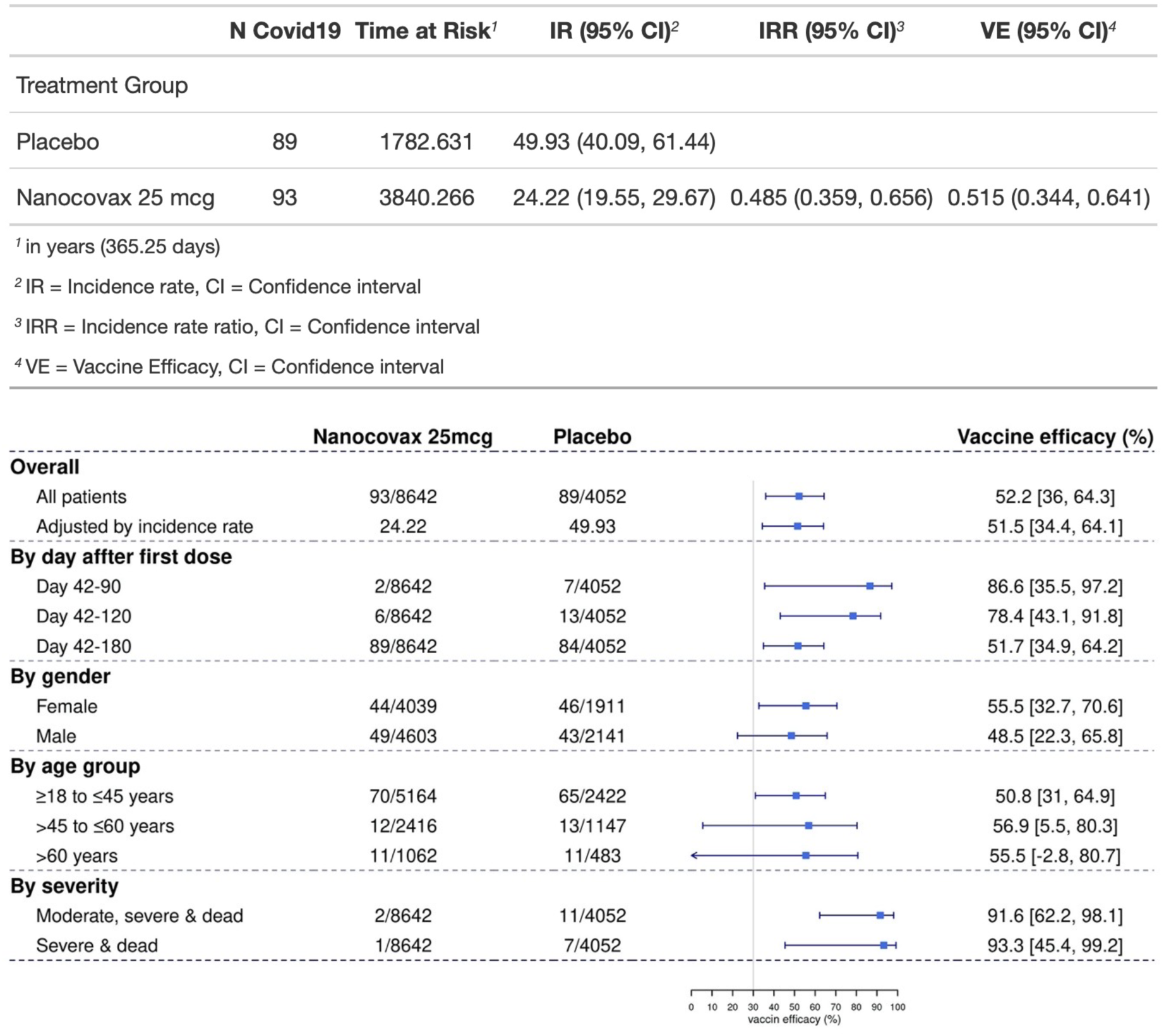
Vaccine efficacy of Nanocovax in preventing Covid-19 by subgroups and timeframes, based on per-protocol population. The VE was defined as 1-the hazard ratio(vaccine/placebo), and 95% CI were calculated using stratified Cox proportional hazards model with Efron’s method of tie handling and treatment groups as a covariate.

The protection against symptomatic infection was waned over time. The VE were 86.6% [35.5-97.2], 78.4% [43.1-91.8] and 51.7% [34.9-64.2] at 3 months, 4 months, and 6 months after the first dose, respectively. Female participants appeared to have higher protection (55.5% [32.7-70.6]), compared to male participants 48.5% [22.3-65.8]. The youngest age group of 18 to 45 years old appeared to have lower protection 50.8% [31.0-64.9] than older age groups, over 45 to 60 and over 60: 55.9% [5.5-80.3] and 55.5% [-2.8-80.7], respectively. However, the lower bound VE of 18-45 age group was 31% which was higher than 5.5% and -2.8% of over 45 to 60, and over 60 years old groups. This was likely due to the small numbers of Covid-19 cases observed in these groups. Remarkably, the protection against severe illness/death was 93.3% [45.4-99.2]) (Figure 6).

## 4. Discussion

The VE of Nanocovax against symptomatic infection in Phase 2 and 3 were 86.6% [35.6-97.2], 78.4% [43.1-91.8] and 51.7 [34.9-63.4] for the periods of 3 months, 4 months and 6 months after the first vaccination, respectively. The overall combined VE against symptomatic infection in Phase 2 and 3 was 51.5% [34.4-64.1]. Importantly, the VE against severe illness and death was 93.3% [45.4-99.2]. Although the lower bound of the VE confidence interval was below 50%, it met the FDA guidance for Covid-19 vaccine development that the efficacy should be at least 50% and lower bound of confidence interval around primary efficacy endpoint estimate to be higher than 30%^6^.

The VE analysis of Nanocovax was complicated by a number of factors. Firstly, the dominant virus strain changed in Vietnam during the study. Although the trial was designed to evaluate the protection against the original (Wuhan) strain, the Delta variant was the overwhelmingly dominant strain at study sites (Vietnam). In fact, the lower than estimated VE was expected, as several studies on approved vaccines reported a dramatic reduction VE against the Delta strain^7–9^. Secondly, the strict social distancing measures were imposed by the local (Vietnam) government during the first 3 months of the study, making VE analysis during this time frame challenging due to the limited number of Covid-19 cases. Because the efficacy/effectiveness decreases over time as shown by several studies on different Covid-19 vaccines ^10–12^, the missing early efficacy window might have affected the overall efficacy observed during this study. Thirdly, the public availablity of commercially available Covid-19 antibody tests allowed some participants to guess the treatment they had received which resulted in them opting out of the trial if they were sero-negative. Fourthly, participants were not protected from the vaccine mandate issued by the local government. As a result, a number of participants were forced to receive commercially approved vaccines and therefore forfeited their participation in study.

The results of the Phase 3 study demonstrated the excellent safety of Nanocovax. Most recorded adverse events were Grade 1 which disappeared within 48 hours after injection. In comparison to similar studies of approved vaccines^13–17^, Nanocovax appears to have equal or lower incidence of reactogenicity, in terms of local and systemic AE.

Nanocovax induces robust humoral immune responses, in term of S-specific antibody and neutralizing antibody levels. The GFMR of Anti-S IgG on Day 42 was more than 218 compared to the baseline level on Day 0. Neutralizing antibody titers in the Phase 3 study were compared to Phase 1 and Phase 2 (on the Wuhan strain). We observed a drop of neutralizing antibody titers against the Alpha and Delta variants (Figure S5) which was consistent with published studies^25^,which was translated into a decreased VE in this trial in which Delta was the dominant variant.

Nanocovax was found to induce both Th1 and Th2 responses with a polarization to Th2. The Th2 polarization poses a concern of antibody-dependent enhancement (ADE)^6,26^. This concern has been partially addressed with vaccine studies of hamster models challenged with SARS-CoV-2 as well as the approval of therapeutic antibodies for Covid-19 treatment^27,28^. Importantly, ADE was not observed among patients in the vaccine group of this Nanocovax trial.

This Phase 3 trial has a number of limitations. As mentioned above, the VE analysis encountered multiple challenges: the emergence of Delta variant, the government polices (strict quarantine, vaccine mandate regardless of vaccine trial enrollment) and the difference in Covid-19 survaillance at different study sites. Other limitations are low ethnic diversity (all Vietnamese), small study sized and limited follow-up time. In fact, due to ethical concerns, especially the safety of all participants, the principal investigators, the sponsor, and Ethics Committee decided to terminate the study in December 2022 (1 year of ahead of the projected end date). All participants received emergency use authorized vaccines after this date.

In conclusion, Nanocovax has excellent safety and immunogenicity profile and robust efficacy.

## Data Availability

All data produced in the present study are available upon reasonable request to the authors

---

**Figure S1.**
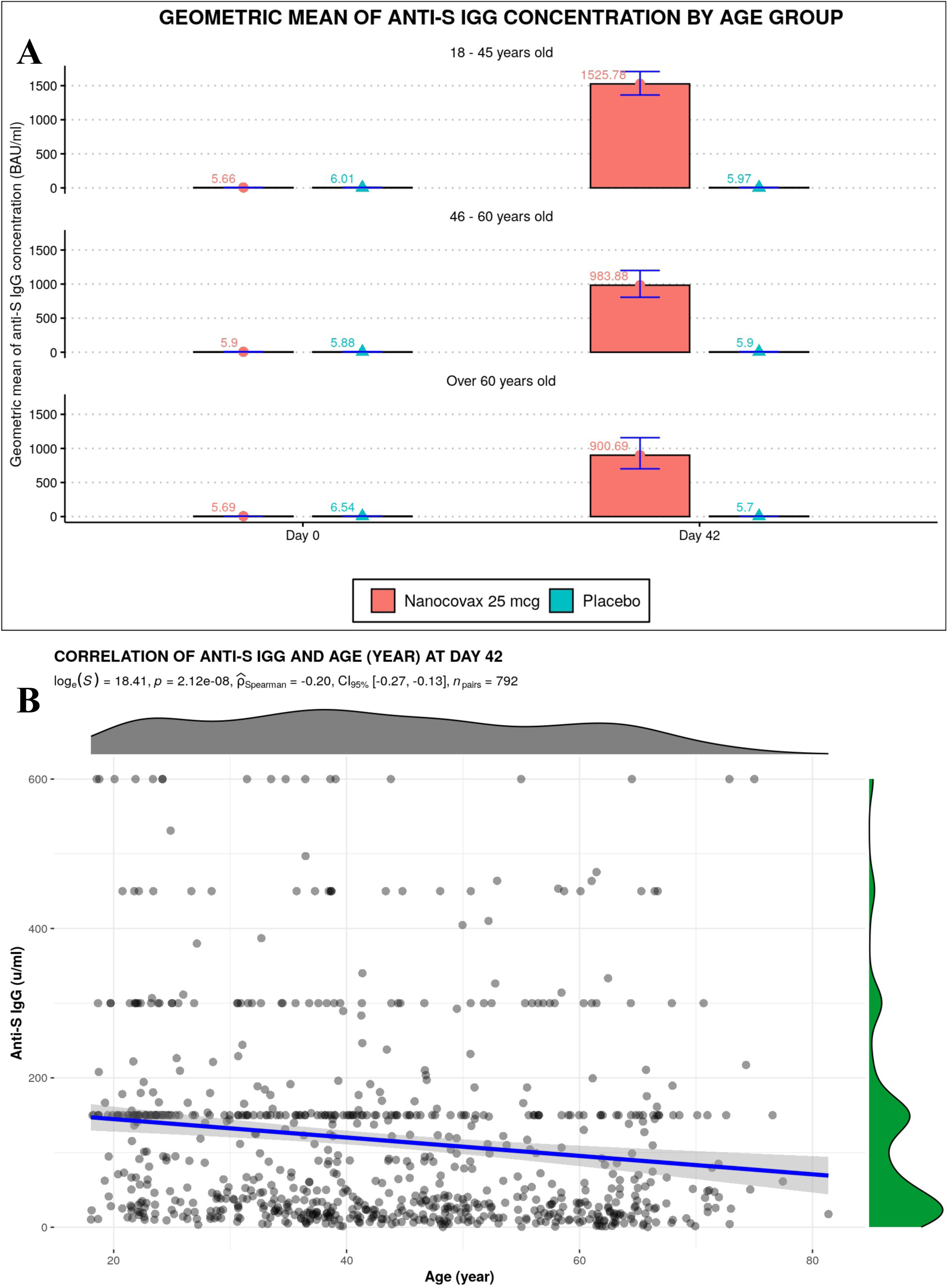
A) Anti-S IgG responses of different age groups. B) Correlation between age and IgG response.

**Figure S2.**
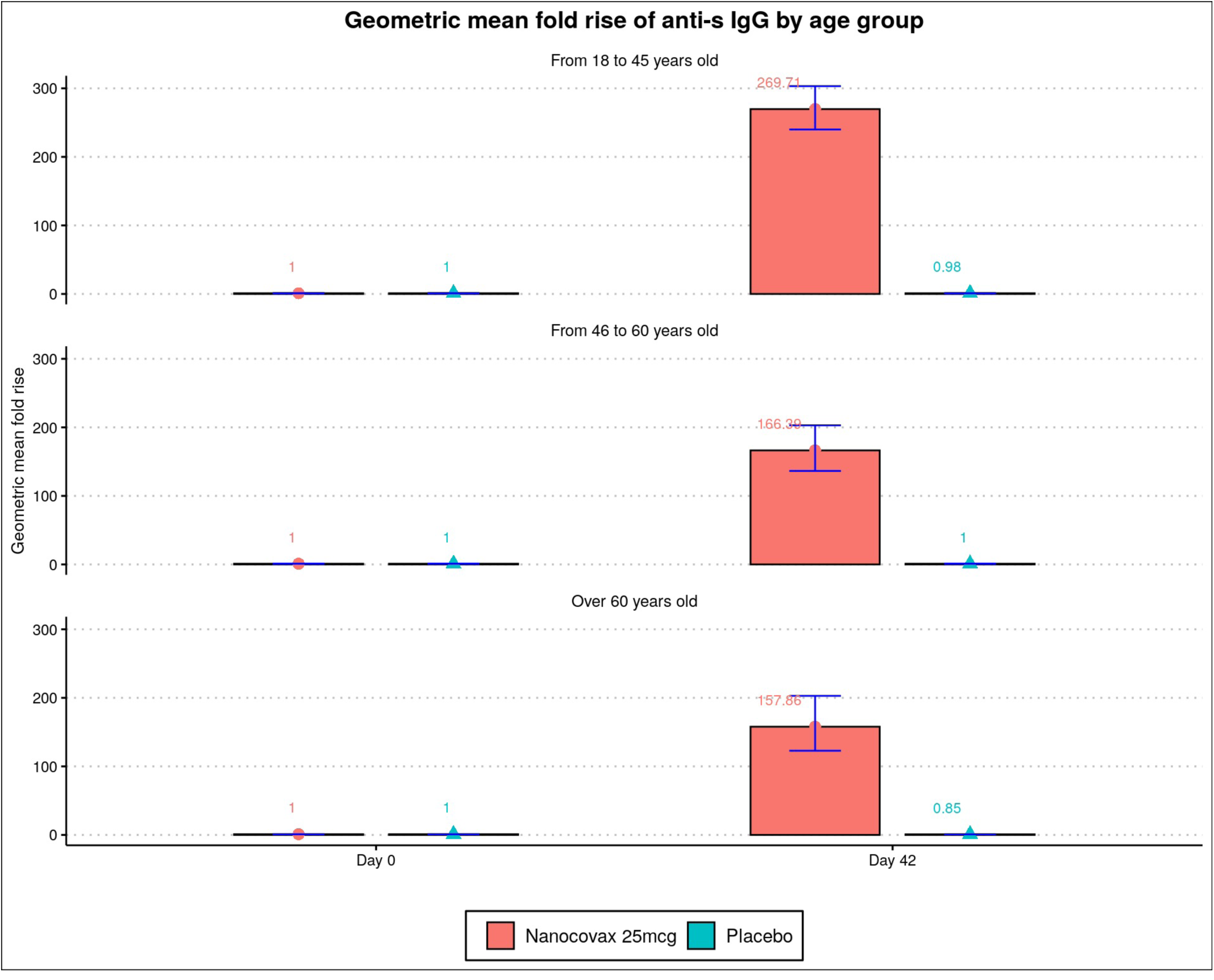
Geometric fold rise of anti-S IgG of different age group.

**Figure S3.**
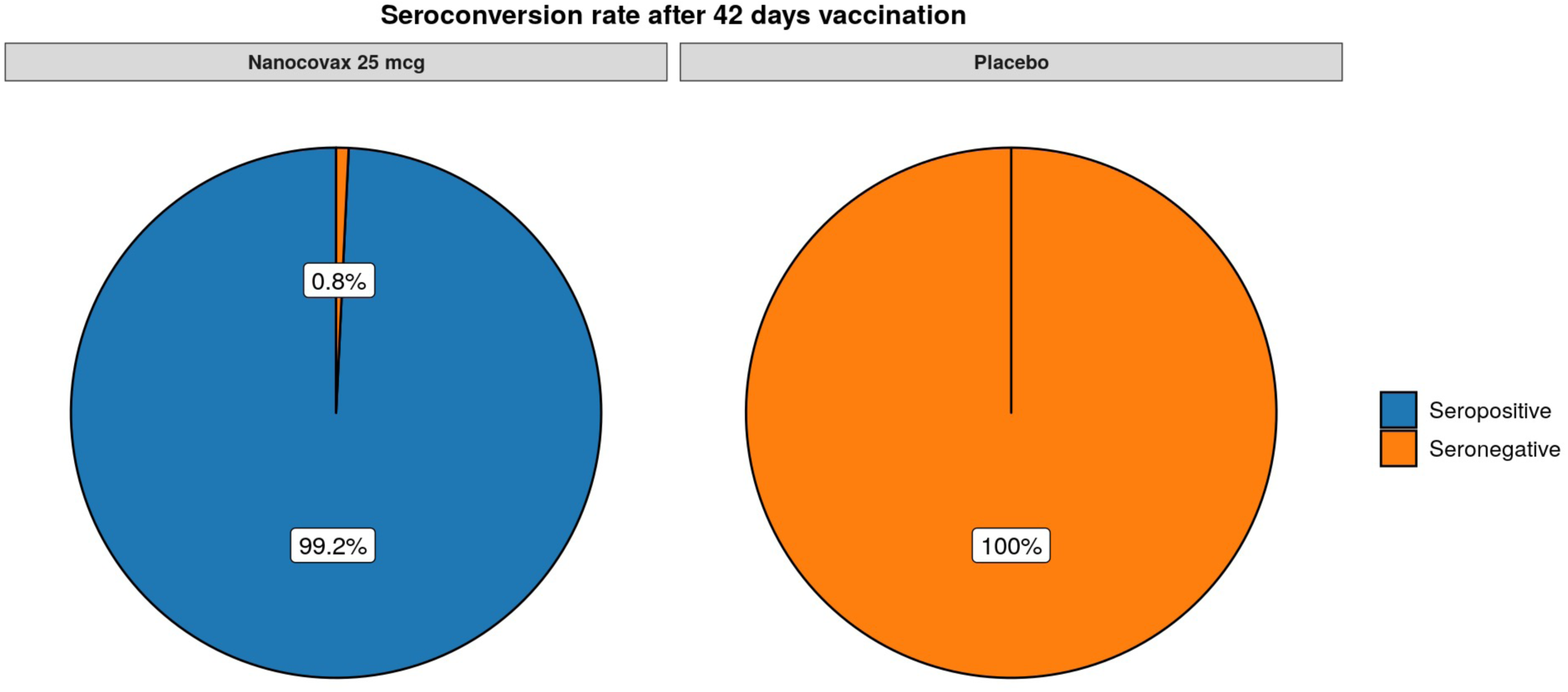
Seroconversion rates of vaccine (n =) and placebo group (n=).

**Figure S4.**
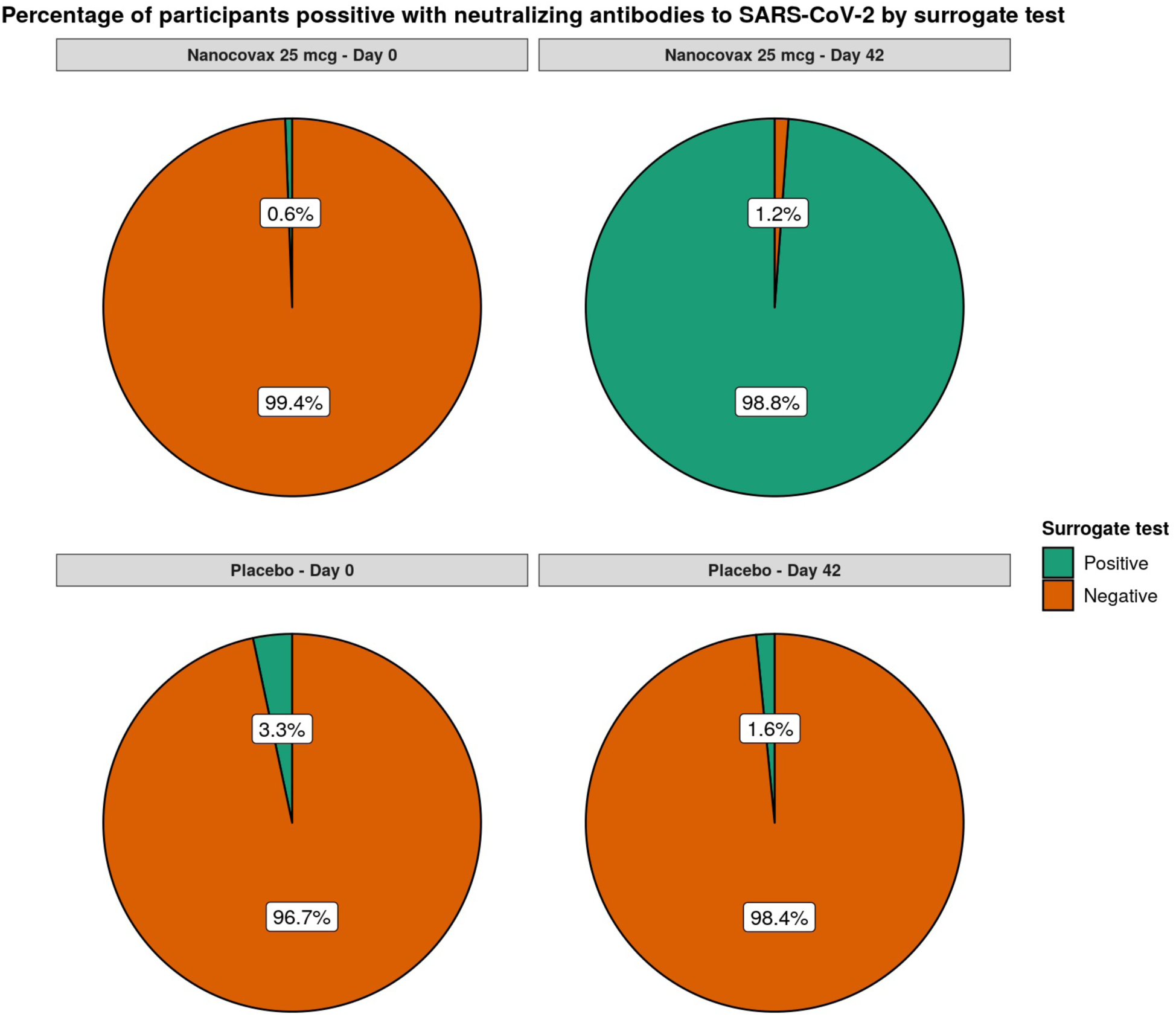
Percentage of participants in vaccine (n= 859) and placebo (n=145) group positive for neutralizing antibody evaluated by surrogate virus neutralization test (sVNT).

**Figure S5.**
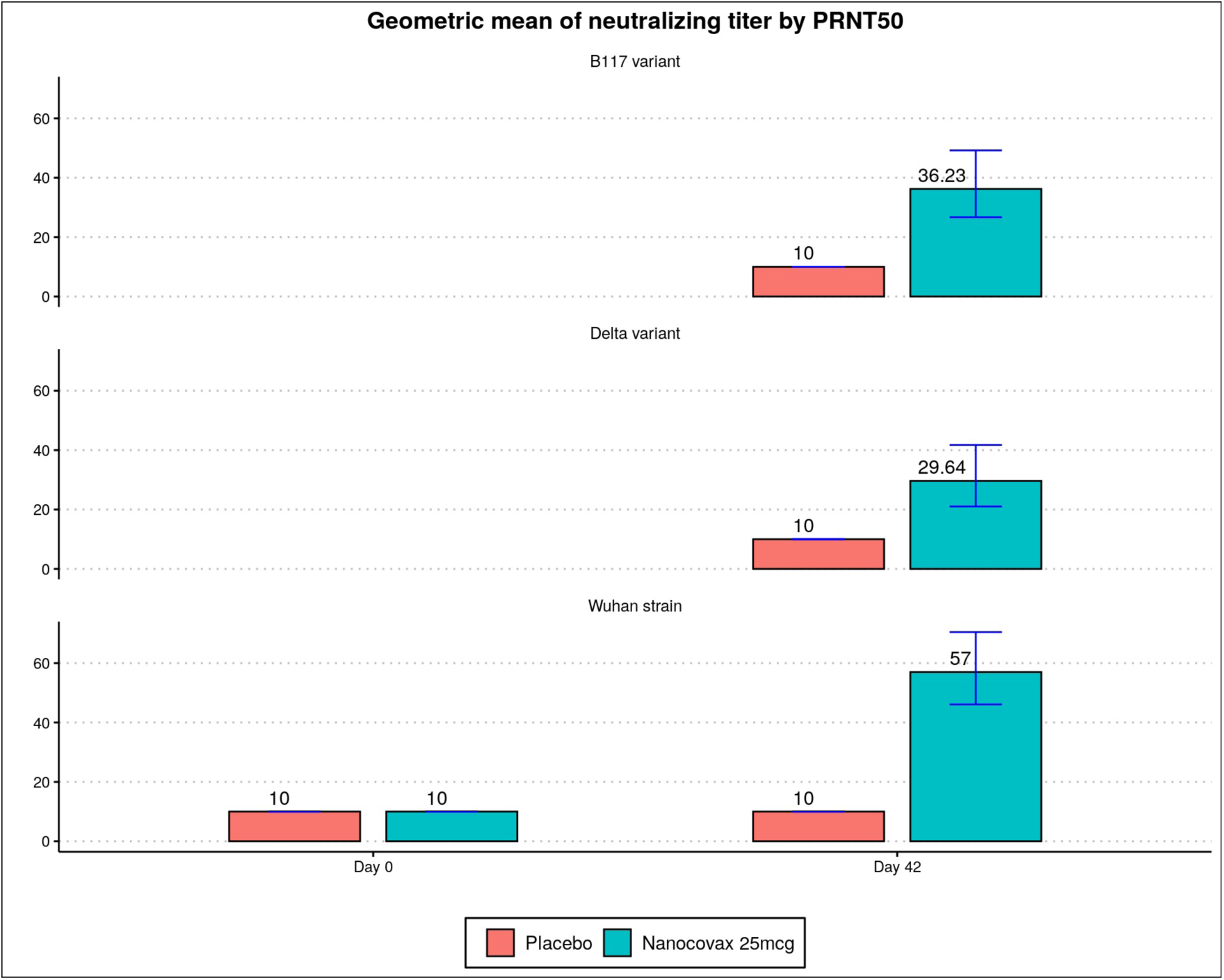
Neutralizing antibody titers (geometric mean) on Wuhan strain B.1.1.7 (Alpha) and B.1.617.2 (Delta) variants, evaluated by PRNT50.

**Figure S6.**
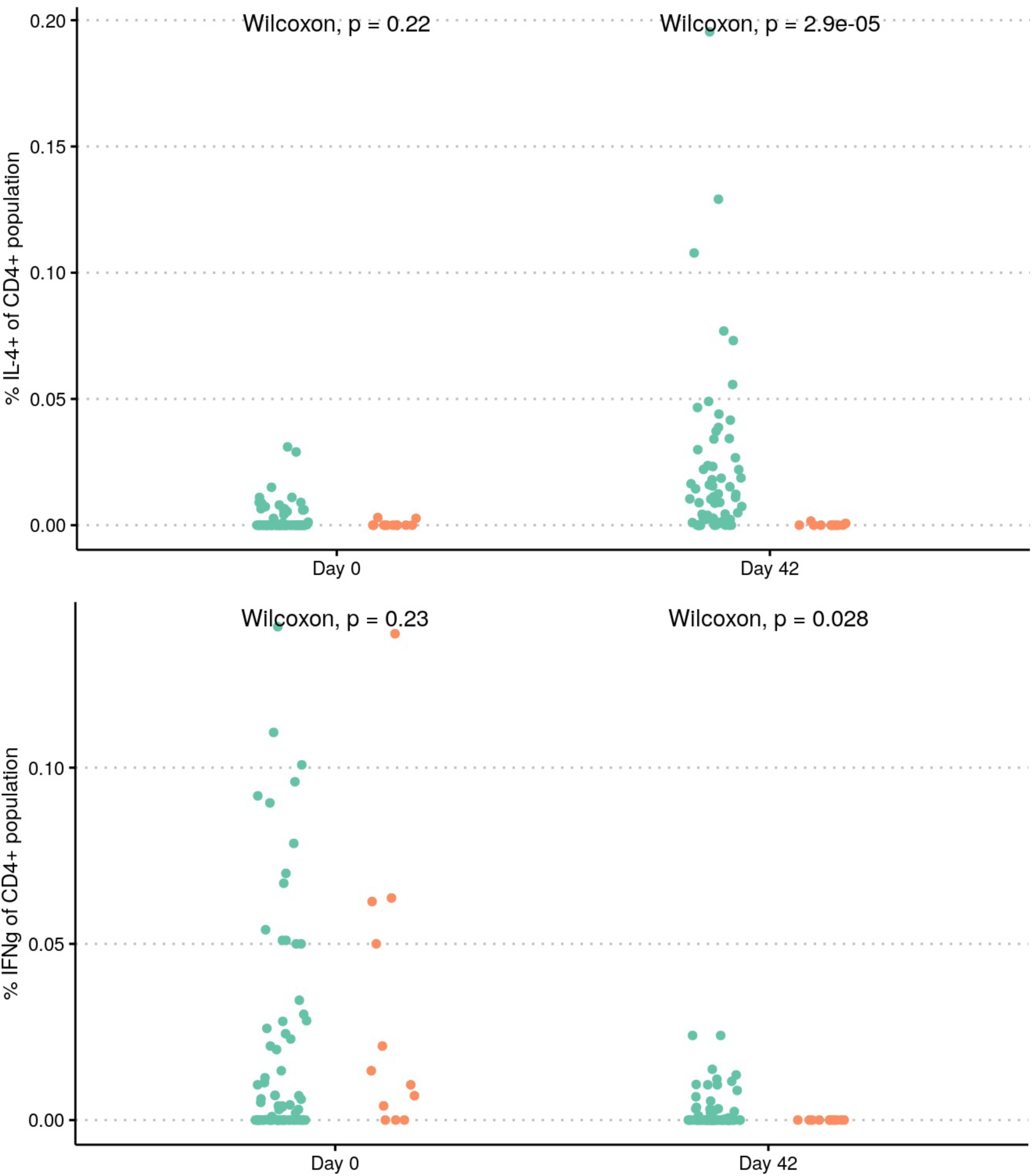
S specific CD4+ T cell responses of Nanocovax. Frequencies of S-specific CD4+ T cell producing IFN-g (Th1) and IL-4 (Th2) on Day 0 (baseline) and on Day 42 (2 weeks after the second dose) of the vaccine (n=66) and placebo (n=11) groups.

**Table S1.** Key trial timing of Phase 3

| Timepoint | 1<sup>st</sup> injection<br>(Day 0) | 2<sup>nd</sup> injection<br>(Day 28) | Day 42 | 6 months | 1 years |
| --- | --- | --- | --- | --- | --- |
| Visit number | 1 | 2 | 3 | 4 | 5 |
| Assessment of immunogenicity<br>(subgroup of 1007 participants) | X | X | X | X | X |
| Assessment of primary endpoint<br>efficacy |  |  | Starting from Day 42 |  |  |
| Assessment of safety and<br>secondary endpoint | Throughout the study |  |  |  |  |

**Table S2.** Clinical background of participants at baseline

| Coexisting condition | Placebo (n = 4,143) |  | Nanocovax (n = 8,864) |  | Total (n = 13,007) |  |
| --- | --- | --- | --- | --- | --- | --- |
|  | n | % | n | % | n | % |
| Chronic liver disease | 14 | 0.34 | 13 | 0.15 | 27 | 0.21 |
| Chronic kidney disease | 5 | 0.12 | 7 | 0.08 | 12 | 0.09 |
| Chronic heart disease | 10 | 0.24 | 12 | 0.14 | 22 | 0.17 |
| Cancer | 2 | 0.05 | 3 | 0.03 | 5 | 0.04 |
| History of allergies | 158 | 3.81 | 427 | 7.82 | 585 | 4.5 |
| History of blood clotting<br>disorder | 2 | 0.05 | 1 | 0.01 | 3 | 0.02 |
| High blood pressure | 133 | 3.21 | 269 | 3.03 | 402 | 3.09 |
| Diabetes | 39 | 0.94 | 88 | 0.99 | 127 | 0.98 |
| Diagnosed with hepatitis B | 18 | 0.43 | 42 | 0.47 | 60 | 0.46 |
| Diagnosed with hepatitis C | 0 | 0 | 0 | 0 | 0 | 0 |

**Table S3.** Solicited local adverse events within 7 days of the first and second injections.

| After 1st injection |  | Placebo<br>(n=3,956) | Nanocovax 25 mcg<br>(n=8,515) | Total<br>(n=12,456) |
| --- | --- | --- | --- | --- |
| Pain | Not observed | 2,375 (60,0%) | 5,057 (59.4%) | 7,433 (59.6%) |
|  | Mild | 1,515 (38.3%) | 3,340 (39.2%) | 4,855 (38.9%) |
|  | Moderate | 59 (1.5%) | 111 (1.3%) | 170 (1.4%) |
|  | Severe | 7 (0.2%) | 7 (0.1%) | 14 (0.1%) |
| Redness | Not observed | 3,920 (99,1%) | 8,440 (99.1%) | 12,360 (99.1%) |
|  | Mild | 33 (0.8%) | 66 (0.8%) | 99 (0.8%) |
|  | Moderate | 3 (0.1%) | 8 (0.1%) | 11 (0.1%) |
|  | Severe | 0 (0.0%) | 1 (0.0%) | 1 (0.0%) |
| Itching | Not observed | 3,755 (94,9%) | 8,112 (95.3%) | 11,867 (95.2%) |
|  | Mild | 198 (5.0%) | 400 (4.7%) | 598 (4.8%) |
|  | Moderate | 3 (0.1%) | 3 (0.0%) | 6 (0.0%) |
| Tenderness | Not observed | 2,928 (74%) | 6,240 (73.3%) | 9,168 (73.5%) |
|  | Mild | 754 (19.1%) | 1,633 (19.2%) | 2,387 (19.1%) |
|  | Moderate | 271 (6.9%) | 628 (7.4%) | 899 (7.2%) |
|  | Severe | 3 (0.1%) | 14 (0.2%) | 17 (0.1%) |
| Swelling/firmness | Not observed | 3,924 (99,2%) | 8,451 (99.2%) | 12,375 (99.2%) |
|  | Mild | 29 (0.7%) | 55 (0.6%) | 84 (0.7%) |
|  | Moderate | 3 (0.1%) | 7 (0.1%) | 10 (0.1%) |
|  | Missing | 0 (0.0%) | 2 (0.0%) | 2 (0.0%) |
| After 2nd injection |  | Placebo<br>(n=2,439) | Nanocovax 25 mcg<br>(n=5,349) | Total<br>(n=7,788) |
| Pain | Not observed | 1,778 (72.9%) | 3,763 (70.3%) | 5,541 (71.1%) |
|  | Mild | 646 (26.5%) | 1,536 (28.7%) | 2,182 (28.0%) |
|  | Moderate | 15 (0.6%) | 49 (0.9%) | 64 (0.8%) |
|  | Severe | 0 (0%) | 1 (0%) | 1 (0%) |
| Redness | Not observed | 2,435 (99.8%) | 5,320 (99.5%) | 7,755 (99.6%) |
|  | Mild | 4 (0.2%) | 24 (0.4%) | 28 (0.4%) |
|  | Moderate | 0 (0%) | 3 (0.1%) | 3 (0%) |

|  |  |  |  |  |
| --- | --- | --- | --- | --- |
|  | Severe | 0 (0%) | 2 (0%) | 2 (0%) |
| **Itching** | Not observed | 2,359 (96.7%) | 5,077 (94.9%) | 7,436 (95.5%) |
|  | Mild | 78 (3.2%) | 265 (5.0%) | 343 (4.4%) |
|  | Moderate | 2 (0.1%) | 7 (0.1%) | 9 (0.1%) |
| **Tenderness** | Not observed | 2,068 (84.8%) | 4,386 (82.0%) | 6,454 (82.9%) |
|  | Mild | 293 (12.0%) | 753 (14.1%) | 1,046 (13.4%) |
|  | Moderate | 77 (3.2%) | 203 (3.8%) | 280 (3.6%) |
|  | Severe | 1 (0%) | 7 (0.1%) | 8 (0.1%) |
| **Swelling/firmness** | Not observed | 2,431 (99.7%) | 5,318 (99.4%) | 7,749 (99.5%) |
|  | Mild | 7 (0.3%) | 26 (0.5%) | 33 (0.4%) |
|  | Moderate | 1 (0%) | 2 (0%) | 3 (0%) |
|  | Severe | 0 (0%) | 3 (0.1%) | 3 (0%) |

**Table S4.** Solicited systemic adverse events within 7 days of the first and second injections.

| After 1st injection |  | Placebo<br>(n=3,957) | Nanocovax 25 mcg<br>(n=8,515) | Total<br>(n=12,472) |
| --- | --- | --- | --- | --- |
| Vomiting/Nausea | Not observed | 3,845 (97.2%) | 8,262 (97.0%) | 12,107 (97.1%) |
|  | Mild | 101 (2.6%) | 234 (2.8%) | 335 (2.7%) |
|  | Moderate | 9 (0.2%) | 18 (0.2%) | 27 (0.2%) |
|  | Severe | 1 (0.0%) | 1 (0.0%) | 2 (0.0%) |
| Myalgia | Not observed | 3,258 (82.3%) | 7,153 (84.0%) | 10,411 (83.5%) |
|  | Mild | 656 (16.6%) | 1,277 (15.0%) | 1,933 (15.5%) |
|  | Moderate | 36 (0.9%) | 74 (0.9%) | 110 (0.9%) |
|  | Severe | 6 (0.2%) | 9 (0.1%) | 15 (0.1%) |
|  | Missing | 0 (0.0%) | 2 (0.0%) | 2 (0.0%) |
| Headache | Not observed | 3,297 (83.3%) | 7,133 (83.8%) | 10,430 (83.6%) |
|  | Mild | 599 (15.1%) | 1,265 (14.8%) | 1,864 (14.9%) |
|  | Moderate | 56 (1.4%) | 107 (1.3%) | 162 (1.3%) |
|  | Severe | 5 (0.1%) | 10 (0.1%) | 15 (0.1%) |
| Arthralgia | Not observed | 3,528 (89.1%) | 7,649 (89.8%) | 11,177 (89.6%) |
|  | Mild | 397 (10.1%) | 806 (9.5%) | 1,203 (9.6%) |
|  | Moderate | 25 (0.6%) | 51 (0.6%) | 76 (0.6%) |
|  | Severe | 6 (0.2%) | 7 (0.1%) | 13 (0.1%) |
|  | Missing | 0 (0.0%) | 2 (0.0%) | 2 (0.0%) |
| Fatigue | Not observed | 2,998 (75.7%) | 6,507 (76.4%) | 9,505 (76.2%) |
|  | Mild | 898 (22.7%) | 1,890 (22.2%) | 2,788 (22.3%) |
|  | Moderate | 55 (1.4%) | 112 (1.3%) | 167 (1.3%) |
|  | Severe | 4 (0.2%) | 6 (0.1%) | 10 (0.1%) |
|  | Very severe | 1 (0.0%) | 0 (0.0%) | 1 (0.0%) |

|  |  |  |  |  |
| --- | --- | --- | --- | --- |
| **Chill** | Not observed | 3,585 (90.6%) | 7,778 (91.3%) | 11,363 (91.1%) |
|  | Mild | 350 (8.8%) | 700 (8.2%) | 1050 (8.4%) |
|  | Moderate | 19 (0.5%) | 33 (0.4%) | 52 (0.4%) |
|  | Severe | 2 (0.0%) | 2 (0.0%) | 3 (0.0%) |
|  | Very severe | 1 (0.0%) | 0 (0.0%) | 1 (0.0%) |
|  | Missing | 0 (0.0%) | 2 (0.0%) | 2 (0.0%) |
| **Fever** | Not observed | 3,827 (96.7%) | 8,258 (97.0%) | 12,085 (96.9%) |
|  | Mild | 119 (3.0%) | 243 (2.8%) | 362 (2.9%) |
|  | Moderate | 8 (0.2%) | 12 (0.1%) | 20 (0.2%) |
|  | Severe | 2 (0.1%) | 2 (0.0%) | 4 (0.0%) |
| **Diarrhea** | Not observed | 3,829 (96.8%) | 8,251 (96.9%) | 12,080 (96.9%) |
|  | Mild | 117 (3.0%) | 249 (2.9%) | 366 (2.9%) |
|  | Moderate | 9 (0.2%) | 15 (0.2%) | 24 (0.2%) |
|  | Severe | 1 (0.0%) | 0 (0.0%) | 1 (0.0%) |
| **After 2nd injection** |  | **Placebo**<br>**(n=2,439)** | **Nanocovax 25 mcg**<br>**(n=5,349)** | **Total**<br>**(n=7,788)** |
| **Vomiting/Nausea** | Not observed | 2,413 (98.9%) | 5,278 (98.7%) | 7,691 (98.8%) |
|  | Mild | 24 (1.0%) | 68 (1.3%) | 92 (1.2%) |
|  | Moderate | 1 (0.0%) | 3 (0.1%) | 4 (0.1%) |
|  | Severe | 1 (0.0%) | 0 (0.0%) | 1 (0.0%) |
| **Myalgia** | Not observed | 2,218 (90.9%) | 4,827 (90.2%) | 7,045 (90.5%) |
|  | Mild | 215 (8.8%) | 487 (9.1%) | 702 (9.0%) |
|  | Moderate | 3 (0.1%) | 33 (0.6%) | 36 (0.5%) |
|  | Severe | 3 (0.1%) | 2 (0.0%) | 5 (0.1%) |
| **Headache** | Not observed | 2,258 (92.6%) | 4,929 (92.1%) | 7,187 (92.3%) |

|  |  |  |  |  |
| --- | --- | --- | --- | --- |
|  | Mild | 167 (6.8%) | 390 (7.3%) | 557 (7.2%) |
|  | Moderate | 9 (0.4%) | 27 (0.5%) | 36 (0.5%) |
|  | Severe | 5 (0.2%) | 3 (0.1%) | 8 (0.1%) |
| **Arthralgia** | Not observed | 2,312 (94.8%) | 5,006 (93.6%) | 7,318 (94.0%) |
|  | Mild | 119 (4.9%) | 319 (6.0%) | 438 (5.6%) |
|  | Moderate | 6 (0.2%) | 22 (0.4%) | 28 (0.4%) |
|  | Severe | 2 (0.1%) | 2 (0.0%) | 4 (0.1%) |
| **Fatigue** | Not observed | 2,104 (86.3%) | 4,538 (84.8%) | 6,642 (85.3%) |
|  | Mild | 317 (13.0%) | 764 (14.3%) | 1,081 (13.9%) |
|  | Moderate | 14 (0.6%) | 44 (0.8%) | 58 (0.7%) |
|  | Severe | 4 (0.2%) | 3 (0.1%) | 7 (0.1%) |
| **Chill** | Not observed | 2,347 (96.2%) | 5,103 (95.4%) | 7,450 (95.7%) |
|  | Mild | 89 (3.6%) | 236 (4.4%) | 325 (4.2%) |
|  | Moderate | 3 (0.1%) | 10 (0.2%) | 13 (0.2%) |
| **Fever** | Not observed | 2,407 (98.7%) | 5,274 (98.6%) | 7,681 (98.6%) |
|  | Mild | 27 (1.1%) | 69 (1.3%) | 96 (1.2%) |
|  | Moderate | 2 (0.1%) | 4 (0.1%) | 6 (0.1%) |
|  | Severe | 2 (0.1%) | 2 (0.0%) | 4 (0.1%) |
|  | Very severe | 1 (0.0%) | 0 (0.0%) | 1 (0.0%) |
| **Diarrhea** | Not observed | 2,386 (97.8%) | 5,240 (98.0%) | 7,626 (97.9%) |
|  | Mild | 47 (1.9%) | 102 (1.9%) | 149 (1.9%) |
|  | Moderate | 6 (0.2%) | 7 (0.1%) | 13 (0.2%) |

**Table S5.** Unsolicited adverse events and their relevance to investigational product.

| **Adverse events** | **Placebo**<br>**(n=4.143)** | **Nanocovax 25 mcg**<br>**(n=8.864)** | **Total**<br>**(n=13.007)** |
| --- | --- | --- | --- |
| **All** | **70 (1.69%)** | **149 (1.68%)** | **219 (1.68%)** |
| Normal | 8 (0.19%) | 14 (0.16%) | 22 (0.17%) |
| Mild | 51 (1.23%) | 98 (1.11%) | 149 (1.15%) |
| Moderate | 8 (0.19%) | 23 (0.26%) | 31 (0.24%) |
| Severe | 3 (0.07%) | 14 (0.16%) | 17 (0.13%) |

