## Supplemental appendix for "The efficacy, safety and immunogenicity Nanocovax: results of a randomized, double-blind, placebo-controlled Phase 3 trial": 4. Supplementary appendix.docx

**1. Vaccination pause rules**

Adverse events meeting any one of the following criteria will result in a hold being placed on

subsequent vaccinations pending further review by the safety monitoring committee (SMC):

• Any SAE attributed to vaccine.

• Any toxicity grade 3 (severe) solicited single AE term occurring in ≥ 7 participants across any vaccine dose following vaccination (first and second vaccinations to be assessed separately).

• Toxicity grade 3 (severe) solicited single prespecified laboratory value occurring in

≥ 7 participants across any vaccine dose following injection (first and second vaccinations to be assessed separately). Prespecified laboratory values to be evaluated include creatinine, alanine aminotransferase, aspartate aminotransferase, bilirubin, hemoglobin, complete white blood count, and platelets.

• Any grade 3 (severe) unsolicited single AE preferred term for which the investigator assesses as related which occurs in ≥ 7 participants across any vaccine dose, within 49 days following vaccination (first).

**2. Realtime RT-PCR testing**

Real-Time Polymerase Chain Reaction (RT-PCR) for detection of SARS-CoV-2 RNA was performed at the study sites, the Vietnam Military Medical Academy and the Pasteur Institute at Ho Chi Minh city). SARS-CoV-2 RNA was isolated by MagMAX^TM^ Viral/Pathogen nucleic acid isolation kit (#A42352). Primers specifically target RdRP and E genes.

**3. Anti-SARS-CoV-2 spike protein serum IgG**

Anti-S IgG in serum samples were evaluated using ADVIA Centaur SARS-CoV-2 IgG (sCOVG) kit (REF# 11207376/11207377) and ADVIA Centaur XP/XPT system by Siemens. All samples were processed according to the manufacturer’s procedures with appropriate controls and calibrators by trained laboratory staff. Results of SARS-CoV-2 IgG are given as Index Unit per ml (U/ml), whereby the cut-off for positivity is defined as ≥ 1·0 U/ml. Limit of detection and limit of quantification are both 0·50 U/ml. The range of quantification is 0·5- 150·0 U/ml, according to manufacturer’s report.

**4. Surrogate virus neutralization assay**

In this assay, the neutralizing activity of anti-S antibody was evaluated by the inhibition of receptor binding domain (RBD) on S protein to its receptor angiotensin-converting enzyme 2 (ACE2) immobilized onto surface of 96 microtiter well plate wells using ELISA based cPass™ SARS-CoV-2 Neutralization Antibody Detection kit (REF # L00847/L00847-5, GenScript). All samples were processed according to the manufacturer’s procedures with specific controls. Results are given as inhibition percentage (%) with the cut-off value of 30%. Samples with ≥ 30% inhibition are considered positive for SARS-CoV-2 neutralizing antibody and negative otherwise.

**5. Plaque reduction neutralization test at dilution reducing more than 50% number of plaque (PRNT_50_)**

All serum samples were heat inactivated at 56^0^C for 30 minutes to remove complement and allowed to equilibrate to room temperature prior to processing for neutralization titer. Samples were diluted in duplicate to an initial dilution of 1:5 followed by 1:2 serial dilutions resulting in a 6-dilution series with each well containing 100 μL. All dilutions were performed in DMEM (Gibco, 11965-092), supplemented with 10% (v/v) fetal bovine serum (heat inactivated, Sigma), 1% (v/v) penicillin/streptomycin (Gibco, 15140-122), and 1% (v/v) L-glutamine (2 mM final concentration, Gibco, 2503-149). Dilution plates were then transported into the BSL-3 laboratory and 100 μL of diluted SARS-CoV-2 inoculum was added to each well to result in a multiplicity of infection (MOI) of 0·01 upon transfer to 12-well titer plates. An untreated, virus-only control and a negative control (media only) were included on every plate. SARS-CoV-2 either Wuhan strain or UK variant was prepared at the concentration of 2·5 PFU/microliter and mixed with diluted serum samples at ratio 1:1 (v/v). The sample/virus mixture was then incubated at 37^0^C (5.0% CO_2_) for 1 hour before transferring to 12-well titer plates with 90% confluent Vero E6 cells. Titer plates were incubated at 37^0^C (5·0% CO_2_) for 5 days. Plates were then fixed and stained for plaque enumeration. Each sample was tested in duplicate. The highest dilution to show at least 50% plaque reduction was reported as the neutralizing titer.

**6. T cell response by intracellular staining for interferon gamma on CD4+ and CD8+ T cells.**

Peripheral blood mononuclear cells (PBMCs) were isolated using Ficoll-Paque Premium (Cytiva, 17544203) and cryopreserved in fetal bovine serum (Gibco, 10438206) containing 10% (v/v) dimethyl sulfoxide (DMSO) (Sigma, D2650). PBMC were rested 8 hours after thawing. Cells with a viability > 85% proceeded to the following assays. PBMCs were cultured in 96-well U-bottom plates at a density of 1x10^6 cells/well and treated with S1 peptide pools (GensScript, RP30020) at concentration of 2 μg/ml or leukocyte activation cocktail including PMA and Ionomycin (positive control) (BD Biosciences, 550583), or medium only (negative control). After incubation at 37^0^C for 18 hours in the presence of BD GolgiPlug™ (BD Biosciences, 555029), cells were labelled for surface markers CD4, CD8 (BD Biosciences, 340443 and 565310). The intracellular cytokines were detected by antibodies specific for T helper 1 (Th1) cytokine IFNg (BD Biosciences, 557718). The samples were processed using a BD FACSCanto II. Data were analyzed using FACSDiva software by BD.

**Appendix I. Procedure of Phase 3**

| **Number of visits** | 1 | 2 | 3 | 4 | 5 | 6 | 7 | 8 | 9 | 10   **(Study end)** *^(c)^* | 11 |
| --- | --- | --- | --- | --- | --- | --- | --- | --- | --- | --- | --- |
| **Description of examination** | Study site | Study site | Phone call^a^ | Study site | Phone call^a^ | Study site^d^  /Phone call^h^ | Phone call^a^ | Study site^d^/Phone call^h^ | Phone call^a^ | Study site^d^/Phone call^h^ | Study site/Home |
| **Month (M)** | M0 | M0 |  | M1 |  |  |  | M6 | M9 | M12 |  |
| **Visit day** | Screening ^b^ | D1 (1st injection) ^b^ | D7 | D28 (2nd injection) | D35 | D42 | Monthly up to 6 months after 1^st^ injection^c^ | D178 ^b^ | Monthly from month 6 to month 12 | D358 ^b^ |  |
| **Window time (day)** | -3 |  | +3 | -3/+7 | +3 | -1/+4 | -2/+7 | -2/+7 | -2/+7 | ±15 |  |
| **Time from the latest injection (day)** | - | 0 |  | 28/0 |  | 14 |  | 150 |  | 330 |  |
| Study consents, medical history and demographic data | X |  |  |  |  |  |  |  |  |  |  |
| General screening for eligibility | X |  |  |  |  |  |  |  |  |  |  |
| HIV, HBV, HCV screening |  | X^d^ |  |  |  |  |  |  |  |  |  |
| Immediate check-up before receiving investigational products (IP) |  | x |  | x |  |  |  |  |  |  |  |
| Clinical examination and vital sign ^1^ | x | x |  | x |  | x^d^ |  | x^d^ |  | x^d^ |  |
| Pregnancy test^2^ | x | x |  | x |  |  |  |  |  |  |  |
| Randomization |  | x |  |  |  |  |  |  |  |  |  |
| ***Administration of investigation products*** |  | x |  | x |  |  |  |  |  |  |  |
| **Efficacy** |  |  |  |  |  |  |  |  |  |  |  |
| Unscheduled medical examination COVID-19. ^3^ |  |  |  |  |  |  |  |  |  |  | x |
| Follow-up for COVID-19 |  | x | x | x | x | x | x | x | x | x |  |
| RT-PCR (nasopharyngeal/throat swab) ^4^ | X(*) | X |  | X(**) |  |  |  |  |  |  | x |
| Anti-SARS COVID-2 antibody ^4^ |  | X |  |  |  |  |  |  |  |  |  |
| **Immunogenicity** |  |  |  |  |  |  |  |  |  |  |  |
| Anti-S IgG ^4^ |  | X^d^ |  |  |  | X^d^ |  | X^d^ |  | X^d^ |  |
| sVNT (neutralizing Ab)^4^ |  | X­^i^ |  |  |  | X^i^ |  |  |  |  |  |
| T cell ^4^ |  | X^e^ |  |  |  | X^e^ |  |  |  |  |  |
| PRNT (neutralizing Ab) ^4^ |  | X^f^ |  |  |  | X^g^ |  |  |  |  |  |
| **Safety** |  |  |  |  |  |  |  |  |  |  |  |
| Reactogenicity reported on electronic diary (7 days after each injection) ^5^ |  | x |  | x |  |  |  |  |  |  |  |
| Evaluation of reactogenicity ^6^ |  | x | x | x | x | x | x | x | x | x |  |
| Concomitant drugs and other vaccines, if any ^7^ |  | x | x | x | x | x | x | x | x | x | x |
| Unsolicited adverse events^8^ |  | x | x | x | x | x | x | x | x | x |  |

1. Vitality assessments are performed both before and after injection of the study product (D1 and D28), before drawing blood
2. Urine pregnancy test
3. Volunteers with symptoms of COVID-19 will be asked to return to the study site within 72 hours or as soon as possible for an unscheduled visit. They will have nasopharyngeal/throat swab for RT-PCR testing and other clinical evaluations (except when after 14 days, the subject has been in close contact with a suspected COVID-19 patient and has had no clinical symptoms). It is important that some symptoms of COVID-19 overlap with systemic adverse reactions expected after injection of the study product (eg, myalgia, headache, fever, and chills). During the first 7 days after injecting IP, when these solicited adverse reactions are common, the investigator should evaluate and decide whether additional nasopharyngeal/throat sampling is necessary (to avoid missed COVID-19 cases).
4. Perform before IP injection.
5. The volunteer will stay at the study site for 60 minutes after receiving the IP, and self-record the reactogenicity into the provided Electronic Diary under the guidance of staff members. After leaving the study site, volunteers will continue to record symptoms, if any, into the Electronic Diary daily, preferably in the evening of the day of injection of the study product and every day of the next 6 days. After 7 days, any reported adverse events should be evaluated by phone or during follow-up visits. For those who are not fully recorded in the e-diary, they will receive reminder calls.
6. Investigators or authorized training personnel will call volunteers to get information of any unsolicited adverse reactions (including any signs and symptoms associated with COVID-19), adverse events and serious adverse events requiring medical attention or leading to study withdrawal, concomitant medications, and any vaccines not included in the study.
7. All drugs associated with or used to treat a serious adverse event or medical adverse event were recorded from the date of screening through the end of the study.
8. The investigator or authorized personnel will call volunteers in a group of 1,000 immunological assessment groups:

- At least once in 7 days after the first injection;
- At least 1 time in 8 to 28 days after each injection;
- At least 1 time from 28 days to 5 months after injection after the 2nd injection;
- At least 1 time every 3 months after injection after injection 1.

a. Investigators will collect information by phone call on a group of 1000 volunteers who (health status, any adverse events). If the volunteer cannot be contacted on the first call, the 2 more attempts will be made. Text message and email will be used if all calling attempts are failed. Family member (whose contact information is provided by the volunteer) will be contacted as the last resource. For the remaining 12,000 subjects, if there are any adverse events or not enough information in the Electronic Diary, a reminder will be called early, or at least monthly. Volunteers can contact research team with provided contact information if they develop symptoms of COVID-19.
b. Screening day and injection day can be combined if feasible. The screening may be repeated several times within 3 days prior to the 1st injection. If volunteers cannot come to the study site due to the COVID-19 pandemic (social distancing, concentrated isolation, home isolation, etc.), researchers will contact by phone (audio/video) instead of on-site visits. The study included all scheduled examination assessments that could be completed remotely, such as adverse event and concomitant medication assessments. Administration of IP are not allowed for home visits unless volunteers cannot come to study site for 2nd injection due to COVID-19 lockdown/quarantine.
c. To be performed over 13,000 volunteers.
d. To be performed on about 1000 volunteers of the immunogenicity assessment group
e. To be performed on 84 volunteers, randomly selected, in the immunological assessment subgroup
f. To be performed on 14 volunteers, randomly selected, in the immunological assessment subgroup
g. To be performed on 168 volunteers, randomly selected, in the immunological assessment subgroup
h. To be performed on over 12,000 volunteers who were not part of the immunological assessment group
i. To be performed on 200 randomly selected volunteers at day 0 and approximately 1000 at day 42 for immunogenicity assessment.

(*) Only be performed when the subject is suspected of having COVID-19 and has epidemiological factors. Up to 5 pooled samples per RT-PCR can be performed. If RT-PCR results (+) will not be included in the study. (**) Only performed when the subject is suspected of being infected with SARS-CoV-2

**Appendix II. Criteria for Covid-19 positive case**

- At least two of the following systemic symptoms: fever (≥38°C), chills, myalgia, headache, sore throat, loss of taste and smell.
- Or at least one of the following respiratory signs/symptoms: cough, shortness of breath or difficulty breathing, or clinical or X-ray evidence of pneumonia.
- And at least one nasopharyngeal swab, nasal swab or saliva sample (or respiratory tract sample, if hospitalized) is positive for SARS-CoV-2 when tested by RT-PCR.

**Appendix III. Criteria for severe Covid-19 case**

- Clinical signs indicating severe systemic illness, respiratory rate ≥30 per minute, heart rate ≥125 bpm, SpO2 ≤93% with room air at sea level or PaO2/FiO2 <300 mmHg;
- Or respiratory failure/ARDS (defined as the need for mechanical or noninvasive ventilation, high-flow oxygen, or ECMO), evidence of shock (systolic blood pressure <90 mmHg, diastolic blood pressure <60 mmHg or require vasopressors)
- Or have severe acute renal, hepatic or neurological dysfunction;
- Or enter intensive care unit or die.

**Appendix IV. Definition of positive case for Covid-19, basing on 2008/QĐ-BYT issued on April 26th, 2001 entitled “Diagnosis and treatment guidance for Covid-19 caused by SARS-CoV-2” (by the Vietnam Ministry of Health).**

**a) Suspected case**

A. An individual has fever and/or acute respiratory infection that cannot be explained by other causes.

B. An individual having any respiratory symptoms AND a history of travel to/to/to/from/from an endemic area* with COVID-19 disease in the 14 days prior to onset of symptoms OR close contact (**) with a suspected or confirmed case of COVID-19 within the 14 days prior to the onset of symptoms.

* Epidemiological areas: are defined as countries and territories with recorded cases of domestic transmission of COVID-19 or where there is an active outbreak in Vietnam according to "Temporary guidance on surveillance and prevention of COVID-19". , against COVID-19” by the Ministry of Health and updated by the Department of Preventive Medicine.

** Close contact:

- Contact at medical facilities, including: direct care of patients with COVID-19; working with healthcare workers with COVID-19; visiting a sick person or staying in the same room as someone who is sick with COVID-19.

- Direct contact within a distance of ≤ 2 meters with a suspected or confirmed case of COVID-19 during the period of illness. Living in the same household as a suspected or confirmed case of COVID-19 during the illness.

- Working in the same working group or in the same room with the confirmed or suspected case during the disease period.

- In the same group: travel, work, play, party, meeting ... with confirmed or suspected cases during the disease period.

- Traveling in the same vehicle (sitting in the same row, in front of or behind two rows of seats) with a suspected or confirmed case of COVID-19 during the illness.

**b) Confirmed case**

Positively result for SARS-CoV-2 by RT-PCR test conducted at any laboratory approved by the Ministry of Health.
